# PUBLIC KNOWLEDGE, BARRIERS AND FACILITATORS TO UK DIETARY GUIDELINE ADHERENCE: A NATIONALLY REPRESENTATIVE SURVEY

**DOI:** 10.64898/2026.04.27.26351827

**Authors:** Alex Griffiths, Kate Austin, Kirstie Cronin, Jamie Matu, Sarah Gregory, Louisa Ells, Oliver M Shannon

## Abstract

**Background:** Adherence to UK dietary guidelines is poor, yet no evidence exists regarding population-level knowledge of these guidelines, or the barriers and facilitators to adherence. This study aimed to characterise knowledge of UK dietary guidelines and perceived barriers and facilitators to adherence in a nationally representative UK sample.

**Methods:** A cross-sectional survey was administered to 1003 adults recruited via Prolific, matched to the UK population by age, sex, and ethnicity. A 22-item knowledge questionnaire assessed awareness of the Eatwell Guide and broader Dietary Reference Values (DRVs), with both strict and liberal scoring applied. Perceived barriers and facilitators to adherence were assessed using custom questionnaire items informed by the COM-B model and TDF framework.

**Results:** Knowledge of Eatwell Guide recommendations was moderate under strict scoring (53.3%) and improved under liberal scoring (72.5%), despite nearly half of participants reporting no familiarity with the Eatwell Guide. Knowledge of broader DRVs was poor using strict scoring (17.9%) but moderate with liberal scoring (58.9%). The most commonly reported barriers were social (e.g. celebrations), environmental (e.g. access to unhealthy foods), and psychological (e.g., mood). The most strongly endorsed facilitators were economic (e.g. cheaper healthy foods) and health-related (e.g. motivated by weight and mental health).

**Conclusions:** These findings suggest that whilst knowledge of UK dietary guidelines is reasonable, individualised behaviour change approaches alone are unlikely to be sufficient. Meaningful population-level improvements will require complementary structural changes to the food environment.

## INTRODUCTION

The UK Government’s healthy eating guidelines are underpinned by Dietary Reference Values (DRVs) which provide nutrient-based recommendations for the general population. These include consuming an average intake of around 50% of total energy from carbohydrates, emphasising high fibre intake (30g/day) and limiting free sugars to no more than 5% of total energy. They also recommend no more than 35% of energy from total fat, including no more than 11% from saturated fat and around 15% total energy from protein. Salt intake is recommended not to exceed 6g per day (OHID, 2016a). These nutrient-based recommendations are visually represented in a public-facing tool called the Eatwell Guide, launched in 2016 as the UK’s principal healthy eating framework (OHID, 2016b). The Eatwell Guide is a food-based model which promotes a balanced diet including at least five portions of fruit and vegetables per day, starchy carbohydrates with an emphasis on wholegrain and high fibre options, and a variety of protein sources including beans, pulses, and fish. It also advises limiting red and processed meat and foods high in fat, salt and sugar, and recommends adequate fluid intake of 6–8 cups per day (OHID, 2016b; Scarborough et al., 2016).

Evidence linking adherence to the Eatwell Guide with health outcomes has begun to emerge in recent years (Shannon et al., 2024). Multiple large UK cohort studies have reported that greater adherence to the Eatwell Guide is associated with reductions in all-cause mortality (Scheelbeek et al., 2020) and gains in life expectancy of up to 10 years (Fadnes et al., 2023). Prospective evidence has also demonstrated that greater Eatwell Guide adherence is associated with favourable changes in markers of adiposity (Griffiths et al., 2025a) and cardiometabolic health (Gregory et al., 2024). However, adherence to the Eatwell Guide was not associated with incident psoriasis risk in a recent large prospective cohort study, suggesting that its benefits may not extend uniformly across all health outcomes (Zhang et al., 2026). Albeit, this growing evidence base suggests that adherence to UK dietary guidelines is associated with meaningful improvements across a range of clinically relevant health outcomes.

Despite these health promoting effects, adherence to UK dietary guidelines remains poor. Data from a large multi-cohort analysis suggest that fewer than 1% of the UK population meet all Eatwell Guide recommendations (nine of which were quantified in this study), with most meeting just three or four (Scheelbeek et al., 2020). In addition, the latest National Diet and Nutrition Survey (NDNS) data confirm that dietary recommendations continue to be widely unmet across the UK population, with only 17% of adults meeting the 5-a-day fruit and vegetable recommendation, 4% meeting the fibre recommendation, and 19% meeting the free sugars recommendation (OHID, 2025). Sociodemographic variation in adherence has also been reported (Griffiths et al., 2025b), highlighting that barriers to following UK dietary guidelines are likely not evenly distributed across the population.

Dietary behaviour is shaped by a complex interplay of individual, social, cultural and environmental factors (Deslippe et al., 2023; Swinburn et al., 2011). Whilst knowledge of dietary recommendations may facilitate better food choices and engagement with nutrition information, behavioural science evidence consistently demonstrates that knowledge is a necessary but insufficient condition for behaviour change (Jezewska-Zychowicz and Plichta, 2022). According to the COM-B model, behaviour is determined by an individual’s capability, motivation, and the opportunity afforded by their social and physical environment (Michie et al., 2011), suggesting that interventions targeting knowledge alone are unlikely to be sufficient at a population level. Nevertheless, knowledge remains a necessary precondition for behaviour change (Worsley, 2002), and to date no published evidence exists regarding general population knowledge of UK dietary guidelines. Quantifying this alongside barriers and facilitators to adherence could help identify specific knowledge gaps, recognise areas of misconception, and inform the design of effective, person-centred behaviour change interventions. The present study therefore aimed to characterise knowledge of UK dietary guidelines and perceived barriers and facilitators to adherence in a nationally representative UK sample.

## METHODS

### Participants

This study received Leeds Beckett Institutional ethics approval (Application ID: 633518). The questionnaire was administered to a nationally representative sample of 1003 adult participants (matched to the UK population based on age, gender and ethnicity) via Prolific, an online crowdsourcing platform that provides access to a pool of verified research participants. Recruitment via Prolific has previously been used to achieve nationally representative samples for assessing nutrition knowledge (Griffiths et al., 2024) and tracking of dietary behaviour (Shannon et al., 2026) in UK adults. All participants received modest remuneration calculated to approximate ∼£10/h on a pro rata basis.

### Questionnaire development

A custom questionnaire was developed via expert consensus to assess knowledge of UK healthy eating guidelines (including both the Eatwell Guide and broader Dietary Reference Values), perceived adherence, and barriers and facilitators to adhering to UK dietary recommendations. Questions were informed by the COM-B model (Michie et al., 2011) and Theoretical Domains Framework (TDF) (Atkins et al., 2017) to identify targets for dietary behaviour change. The questionnaire was pilot tested using a patient and public involvement and engagement (PPIE) approach with members of the public to ensure comprehensibility, and modifications were made to wording and question order accordingly. The final questionnaire was sectioned into five parts: (1) demographics, (2) perceived adherence to UK dietary guidelines, (3) knowledge of UK dietary guidelines, (4) barriers to adherence, and (5) facilitators to adherence. The final questionnaire was built using an online survey tool (Online Surveys, Bristol, UK).

### Calculation of a UK government healthy eating guideline knowledge index

Knowledge of UK dietary guidelines was assessed using two scoring systems applied to a 22-item knowledge questionnaire. Strict scoring awarded one point for each correct answer only, requiring exact identification of the specific DRV or Eatwell Guide messaging. Liberal scoring applied a more lenient approach to questions with specific quantitative recommendations, awarding one point if the participant selected the correct answer or adjacent response options, reflecting general awareness of a recommendation without precise knowledge of the specific value. For example, for the question asking what proportion of the diet should be made up of starchy carbohydrates, the correct answer of one third was awarded one point under strict scoring, whilst liberal scoring additionally awarded one point for adjacent responses of one quarter or one half, reflecting directional awareness that starchy carbohydrates should make up a substantial proportion of the diet without precise knowledge of the specific recommended value. Both scores were calculated separately for Eatwell Guide specific knowledge (14 items) and broader dietary guideline knowledge (8 items) and expressed as a percentage of the maximum possible score for each subscale. Content validity of each question was established through discussion among the research team and review of published Eatwell Guide and Dietary Reference Value guidance.

### Statistical analysis

Descriptive statistics were used to summarise responses to all questionnaire items, including item-level knowledge responses, reported as frequencies and percentages. Overall knowledge scores were summarised as mean percentage correct and standard deviation for both strict and liberal scoring systems, calculated separately for Eatwell Guide specific knowledge and broader dietary guideline (DRV) knowledge. Subgroup comparisons for ordinal variables (perceptions, barriers, and facilitators items) and knowledge scores were conducted using the Mann-Whitney U test. Chi-square tests were used for nominal variables. Subgroups were defined by age (split at closest to the median: under 50 vs 50 and over), sex (male vs female), ethnicity (white vs non-white), and self-reported BMI (healthy weight vs overweight/obesity). Participants with an underweight BMI and non-binary gender were excluded from sub-group analyses due to small sample sizes. All analyses were conducted in R (version 4.4.0).

## RESULTS

### Participant characteristics

This study recruited 1003 participants, with characteristics (age, sex and ethnicity) representative of the UK population based on UK Census data via Prolific. The sample was mostly white, with marginally more female than male participants. Participants were of varied ages and BMI, and of moderate self-defined socio-economic status. Most were not currently using weight loss medication (95.5%) and had not previously used weight loss medication (97.6%).

**Table 1.**
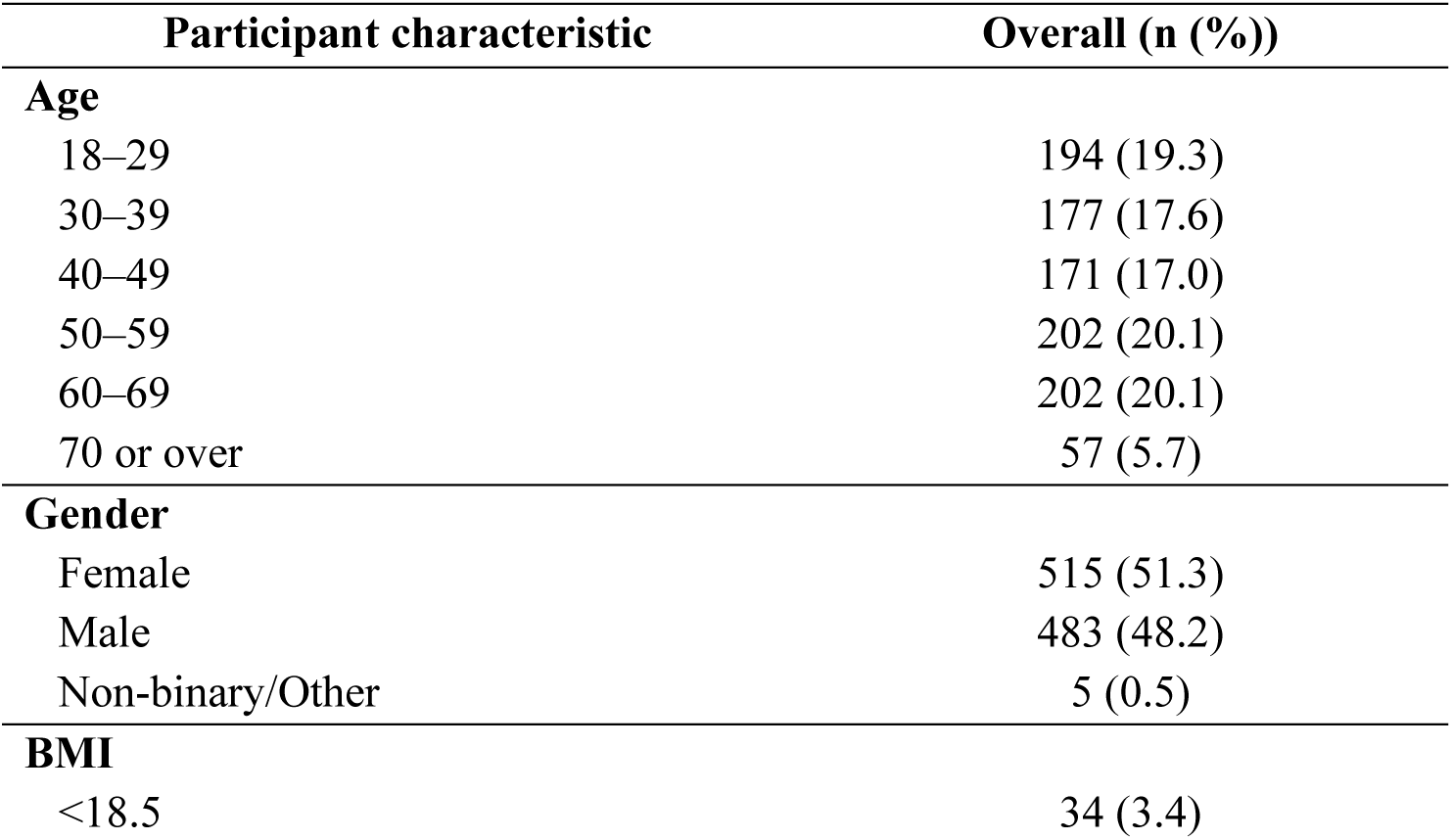
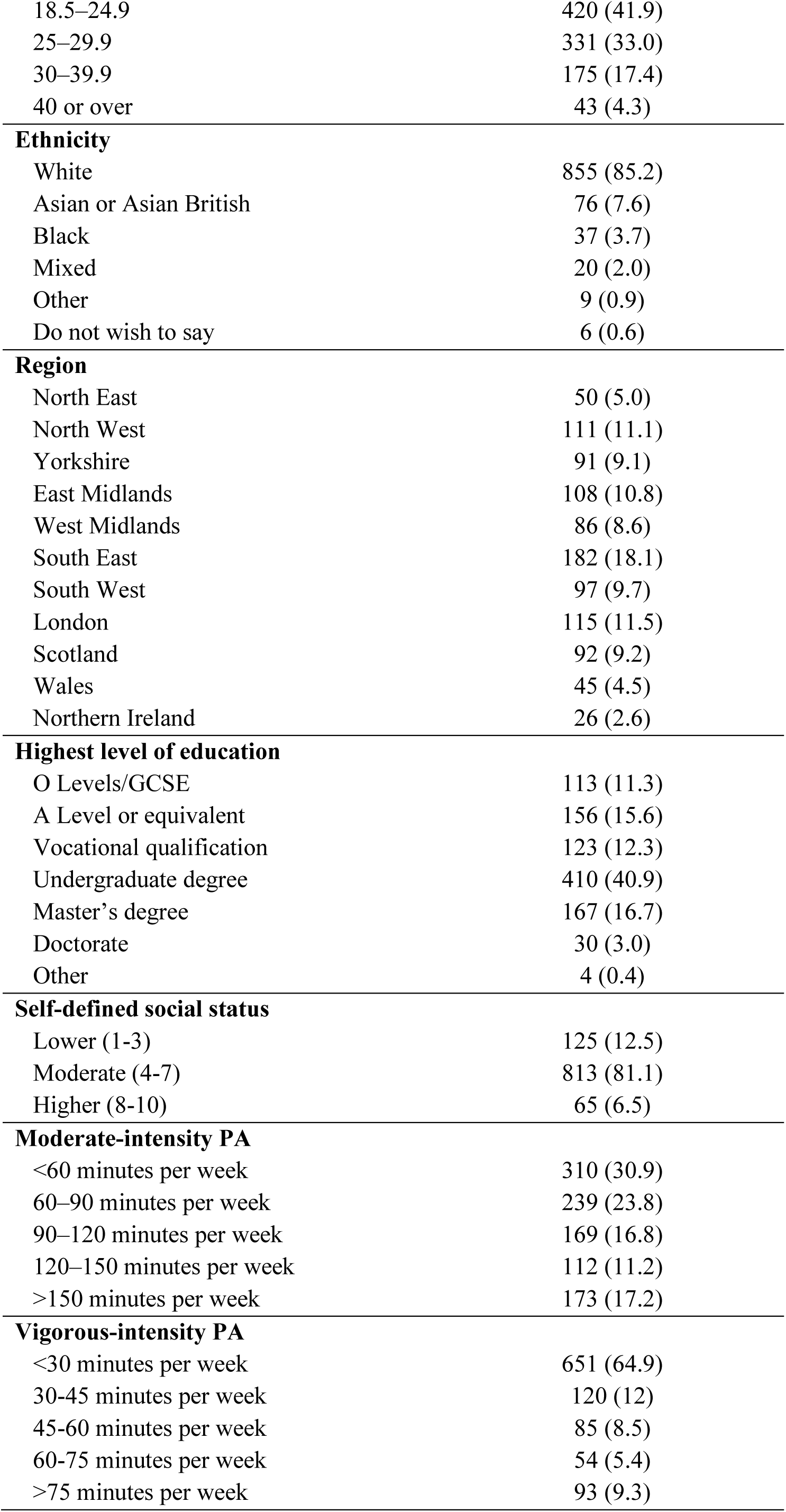
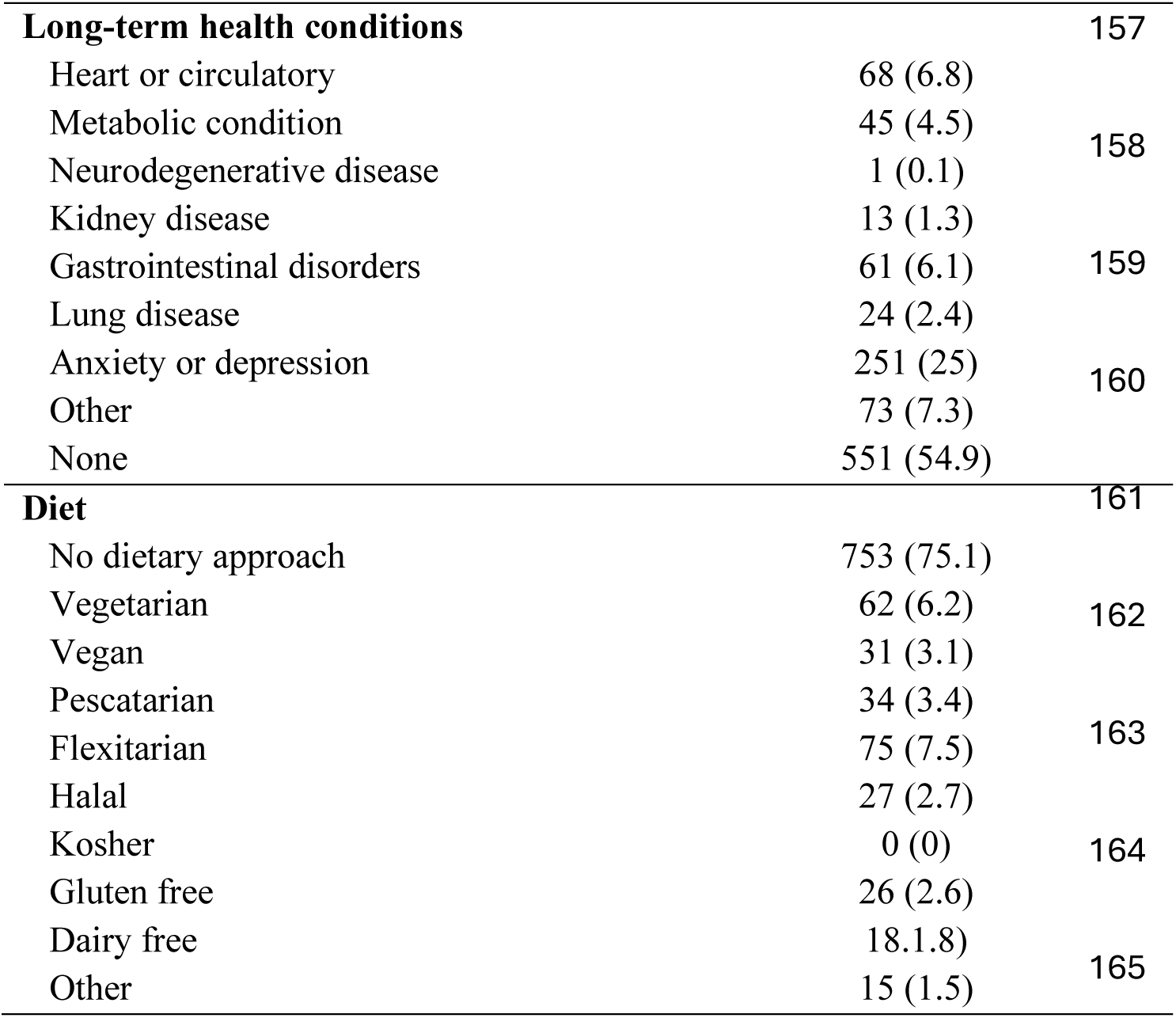
Characteristics of participants.

| Participant characteristic | Overall (n (%)) |
| --- | --- |
| **Age** |  |
| 18–29 | 194 (19.3) |
| 30–39 | 177 (17.6) |
| 40–49 | 171 (17.0) |
| 50–59 | 202 (20.1) |
| 60–69 | 202 (20.1) |
| 70 or over | 57 (5.7) |
| **Gender** |  |
| Female | 515 (51.3) |
| Male | 483 (48.2) |
| Non-binary/Other | 5 (0.5) |
| **BMI** |  |
| <18.5 | 34 (3.4) |
| 18.5–24.9 | 420 (41.9) |
| 25–29.9 | 331 (33.0) |
| 30–39.9 | 175 (17.4) |
| 40 or over | 43 (4.3) |
| **Ethnicity** |  |
| White | 855 (85.2) |
| Asian or Asian British | 76 (7.6) |
| Black | 37 (3.7) |
| Mixed | 20 (2.0) |
| Other | 9 (0.9) |
| Do not wish to say | 6 (0.6) |
| **Region** |  |
| North East | 50 (5.0) |
| North West | 111 (11.1) |
| Yorkshire | 91 (9.1) |
| East Midlands | 108 (10.8) |
| West Midlands | 86 (8.6) |
| South East | 182 (18.1) |
| South West | 97 (9.7) |
| London | 115 (11.5) |
| Scotland | 92 (9.2) |
| Wales | 45 (4.5) |
| Northern Ireland | 26 (2.6) |
| **Highest level of education** |  |
| O Levels/GCSE | 113 (11.3) |
| A Level or equivalent | 156 (15.6) |
| Vocational qualification | 123 (12.3) |
| Undergraduate degree | 410 (40.9) |
| Master's degree | 167 (16.7) |
| Doctorate | 30 (3.0) |
| Other | 4 (0.4) |
| **Self-defined social status** |  |
| Lower (1-3) | 125 (12.5) |
| Moderate (4-7) | 813 (81.1) |
| Higher (8-10) | 65 (6.5) |
| **Moderate-intensity PA** |  |
| <60 minutes per week | 310 (30.9) |
| 60–90 minutes per week | 239 (23.8) |
| 90–120 minutes per week | 169 (16.8) |
| 120–150 minutes per week | 112 (11.2) |
| >150 minutes per week | 173 (17.2) |
| **Vigorous-intensity PA** |  |
| <30 minutes per week | 651 (64.9) |
| 30–45 minutes per week | 120 (12) |
| 45–60 minutes per week | 85 (8.5) |
| 60–75 minutes per week | 54 (5.4) |
| >75 minutes per week | 93 (9.3) |

|  |  |  |
| --- | --- | --- |
| **Long-term health conditions** |  | **157** |
| Heart or circulatory | 68 (6.8) |  |
| Metabolic condition | 45 (4.5) | 158 |
| Neurodegenerative disease | 1 (0.1) |  |
| Kidney disease | 13 (1.3) |  |
| Gastrointestinal disorders | 61 (6.1) | 159 |
| Lung disease | 24 (2.4) |  |
| Anxiety or depression | 251 (25) | 160 |
| Other | 73 (7.3) |  |
| None | 551 (54.9) |  |
| **Diet** |  | **161** |
| No dietary approach | 753 (75.1) |  |
| Vegetarian | 62 (6.2) | 162 |
| Vegan | 31 (3.1) |  |
| Pescatarian | 34 (3.4) |  |
| Flexitarian | 75 (7.5) | 163 |
| Halal | 27 (2.7) |  |
| Kosher | 0 (0) | 164 |
| Gluten free | 26 (2.6) |  |
| Dairy free | 18.1.8) | 165 |
| Other | 15 (1.5) |  |

### Knowledge of UK dietary guidelines

Participants demonstrated moderate knowledge of Eatwell Guide specific recommendations (mean score = 53.3%, SD = 15.4%) and poor knowledge of broader DRVs (mean score = 17.9%, SD = 16.4%) when applying the strict scoring methodology. Under more liberal scoring, Eatwell Guide knowledge improved to 72.5% (SD = 14.0%) and DRV knowledge to 58.9% (SD = 19.2%). Item-level scoring is presented in Supplementary materials 1-2. Notable gaps included identifying fluid types contributing to daily intake (2% strict, 16% liberal) and foods contributing to fruit and vegetable intake (7% strict, 27% liberal). Participants were aware of the types of starchy carbohydrate (95%), and types of protein they should preferentially consume (91%), as well as identifying the number of portions of fruit and vegetables they should consume per day (85%).

### Perceptions of UK dietary guidelines

All data related to perceptions of UK dietary guidelines are presented in Supplementary material 3. Most participants reported being somewhat familiar with UK dietary guidelines (65.2%) however, around half of participants were not at all familiar with the Eatwell Guide (49.3%). Very few participants were aware of culturally adapted versions of the Eatwell Guide (5.7%) or versions adapted for dietary preferences (7.1%). Self-reported adherence to UK dietary guidelines was generally poor, with just 5.6% of participants reporting following guidelines consistently. Despite poor adherence, most participants considered the guidelines moderately or completely relevant to their lives (64.9%) and moderately or very important to follow (69.6%).

Current sources of information about UK dietary guidelines are presented in Supplementary material 4. Notably, 45.1% of participants reported receiving no information about UK dietary guidelines from any source. Government websites (19.2%) and school or education settings (18.3%) were the most commonly reported sources. Using a ranked system (participants scored each source from 1-10), preferred sources of dietary information (Supplementary material 5) were GP or healthcare professionals (mean rank = 3.07, SD = 2.18), followed by government websites (mean rank = 3.51, SD = 2.38) and supermarkets (mean rank = 4.39, SD = 2.50). Notably, despite GP or healthcare professionals being the most preferred source of dietary information, only 14.3% of participants reported currently receiving information from this source.

### Barriers and facilitators to UK dietary guideline adherence

All data related to barriers and facilitators are presented in Table 2. The most endorsed barriers were social (celebrating or attending social events (60.7%)), environmental (availability of unhealthy foods (56.6%)) and psychological (mood (53.2%), comfort eating (40.1%) and difficulty controlling eating (37.2%)). Practical barriers such as cooking tools (5.9%) and finding appropriate foods in shops (8.8%) were among the least endorsed. Cultural barriers appeared low overall but were considerably higher among non-white participants (cultural traditions 32.3%, cultural dietary recommendations 28.1%). Similarly, dietary preferences as a barrier appeared low overall, but was rated more highly as a barrier among those with specific dietary requirements (30.4%).

**Table 2.** Perceived barriers to adhering to UK dietary guidelines, ranked by combined agree percentage.

| Barrier | Strongly Agree n (%) | Agree n (%) | Neither n (%) | Disagree n (%) | Strongly Disagree n (%) | Combined Agree % |
| --- | --- | --- | --- | --- | --- | --- |
| Celebrating with others | 171 (17.0) | 438 (43.7) | 179 (17.8) | 154 (15.4) | 61 (6.1) | 60.7<sup>251</sup> |
| Unhealthy food more available | 173 (17.2) | 395 (39.4) | 176 (17.5) | 177 (17.6) | 82 (8.2) | 56.6 |
| Mood | 144 (14.4) | 389 (38.8) | 155 (15.5) | 210 (20.9) | 105 (10.5) | 53.2<sup>252</sup> |
| Comfort eating | 106 (10.6) | 296 (29.5) | 214 (21.3) | 245 (24.4) | 142 (14.2) | 40.1 |
| Difficulty controlling eating | 72 (7.2) | 301 (30.0) | 145 (14.5) | 301 (30.0) | 184 (18.3) | 37.2<sup>253</sup> |
| Difficult in daily life | 43 (4.3) | 238 (23.7) | 279 (27.8) | 330 (32.9) | 113 (11.3) | 28.0 |
| Expensive | 71 (7.1) | 185 (18.4) | 252 (25.1) | 349 (34.8) | 146 (14.6) | 25.5 |
| Food delivery services | 63 (6.3) | 179 (17.8) | 161 (16.1) | 298 (29.7) | 302 (30.1) | 24.1<sup>254</sup> |
| Estimating portion sizes | 39 (3.9) | 202 (20.1) | 165 (16.5) | 463 (46.2) | 134 (13.4) | 24.0 |
| Time | 49 (4.9) | 154 (15.4) | 178 (17.7) | 407 (40.6) | 215 (21.4) | 20.3<sup>255</sup> |
| Understanding guidelines | 34 (3.4) | 158 (15.8) | 275 (27.4) | 449 (44.8) | 87 (8.7) | 19.2 |
| Nutritional labelling | 28 (2.8) | 158 (15.8) | 153 (15.3) | 433 (43.2) | 231 (23.0) | 18.6 |
| Confidence in meal planning | 28 (2.8) | 136 (13.6) | 134 (13.4) | 429 (42.8) | 276 (27.5) | 16.4<sup>256</sup> |
| Choosing foods in guidelines | 15 (1.5) | 108 (10.8) | 182 (18.1) | 531 (52.9) | 167 (16.7) | 12.3 |
| Cooking skills | 36 (3.6) | 87 (8.7) | 89 (8.9) | 406 (40.5) | 385 (38.4) | 12.3<sup>257</sup> |
| Dietary preferences | 26 (2.6) | 95 (9.5) | 140 (14.0) | 332 (33.1) | 410 (40.9) | 12.1 |
| Cultural traditions | 21 (2.1) | 78 (7.8) | 119 (11.9) | 347 (34.6) | 438 (43.7) | 9.9 |
| Cultural recommendations | 13 (1.3) | 76 (7.6) | 180 (17.9) | 332 (33.1) | 402 (40.1) | 8.9<sup>258</sup> |
| Finding foods in shop | 20 (2.0) | 68 (6.8) | 106 (10.6) | 466 (46.5) | 343 (34.2) | 8.8 |
| Cooking tools | 13 (1.3) | 46 (4.6) | 35 (3.5) | 406 (40.5) | 503 (50.1) | 5.9<sup>259</sup> |
| Vitamins supersede diet | 11 (1.1) | 39 (3.9) | 76 (7.6) | 455 (45.4) | 422 (42.1) | 5.0 |
|  |  |  |  |  |  | 260 |

**Note. Barriers ranked by combined Strongly Agree and Agree percentage (as displayed in the ‘Combined Agree’ column) to show largest to smallest barriers reported. n = 1003.**

Perceived facilitators to adhering to UK dietary guidelines are presented in Table 3. The most commonly endorsed facilitators were economic (healthy foods cheaper (74.0%) and budget meal options (72.1%)) and health related (managing weight (65.8%), improving mental health (65.4%), and improving general health (63.3%)). Portion size examples (61.6%), making healthy foods more enjoyable (61.6%), and healthier options in restaurants and takeaways (60.3%) were also commonly endorsed. Fewer participants endorsed face to face support (40.6%) and ease of understanding the Eatwell Guide (29.7%) as facilitators. Cultural food inclusion appeared low as a facilitator (32.9%), but this was higher among non-white participants (59.1%).

**Table 3.** Perceived facilitators to adhering to UK dietary guidelines, ranked by combined agree percentage.

| Facilitator | Strongly Agree<br>n (%) | Agree n (%) | Neither n (%) | Disagree n (%) | Strongly<br>Disagree n (%) | Combined<br>Agree %<sup>265</sup> |
| --- | --- | --- | --- | --- | --- | --- |
| Healthy foods cheaper | 369 (36.8) | 373 (37.2) | 147 (14.7) | 80 (8.0) | 34 (3.4) | 74.0 |
| Budget meals | 277 (27.6) | 446 (44.5) | 170 (16.9) | 83 (8.3) | 27 (2.7) | 72.1 266 |
| Manage weight | 206 (20.5) | 454 (45.3) | 219 (21.8) | 85 (8.5) | 39 (3.9) | 65.8 |
| Improve mental health | 183 (18.2) | 473 (47.2) | 207 (20.6) | 100 (10.0) | 40 (4.0) | 65.4 267 |
| Improve health | 195 (19.4) | 440 (43.9) | 244 (24.3) | 87 (8.7) | 37 (3.7) | 63.3 |
| Portion size examples | 164 (16.4) | 453 (45.2) | 199 (19.8) | 126 (12.6) | 61 (6.1) | 61.6 |
| Find healthy foods enjoyable | 183 (18.2) | 435 (43.4) | 216 (21.5) | 129 (12.9) | 40 (4.0) | 61.6 268 |
| Restaurants and takeaways<br>offer healthy meals | 211 (21.0) | 394 (39.3) | 209 (20.8) | 130 (13.0) | 59 (5.9) | 60.3 269 |
| Meal plans tailored to dietary<br>preferences | 180 (17.9) | 366 (36.5) | 278 (27.7) | 130 (13.0) | 49 (4.9) | 54.4 |
| App or website support | 124 (12.4) | 415 (41.4) | 241 (24.0) | 149 (14.9) | 74 (7.4) | 53.8 270 |
| Calories on menus | 173 (17.2) | 366 (36.5) | 204 (20.3) | 177 (17.6) | 83 (8.3) | 53.7 |
| Public promotion | 122 (12.2) | 415 (41.4) | 215 (21.4) | 177 (17.6) | 74 (7.4) | 53.6 271 |
| Learning to cook affordable<br>meals | 172 (17.1) | 353 (35.2) | 260 (25.9) | 151 (15.1) | 67 (6.7) | 52.3 |
| Motivated to eat healthily | 105 (10.5) | 413 (41.2) | 315 (31.4) | 127 (12.7) | 43 (4.3) | 51.7 272 |
| Advice tailored to lifestyle | 170 (16.9) | 339 (33.8) | 245 (24.4) | 169 (16.8) | 80 (8.0) | 50.7 |
| Opportunity to learn | 85 (8.5) | 413 (41.2) | 282 (28.1) | 165 (16.5) | 58 (5.8) | 49.7 273 |
| Find EWG easily | 134 (13.4) | 362 (36.1) | 225 (22.4) | 199 (19.8) | 83 (8.3) | 49.5 |
| Family and friends support | 120 (12.0) | 329 (32.8) | 328 (32.7) | 161 (16.1) | 65 (6.5) | 44.8 274 |
| Meal box delivery | 117 (11.7) | 325 (32.4) | 283 (28.2) | 163 (16.3) | 115 (11.5) | 44.1 |
| Workplace healthy options | 125 (12.5) | 289 (28.8) | 312 (31.1) | 161 (16.1) | 116 (11.6) | 41.3 |
| Face to face support | 107 (10.7) | 300 (29.9) | 252 (25.1) | 229 (22.8) | 115 (11.5) | 40.6 275 |
| Cultural foods included | 88 (8.8) | 242 (24.1) | 288 (28.7) | 222 (22.1) | 163 (16.3) | 32.9 |
| EWG easy to understand | 72 (7.2) | 226 (22.5) | 363 (36.2) | 252 (25.1) | 90 (9.0) | 29.7 276 |

**EWG – Eatwell Guide. Note. Items ranked by combined Strongly Agree and Agree percentage (as displayed in the ‘Combined Agree’ column). n = 1003.**

### Sub-group comparisons

Full subgroup data are presented in Supplementary Tables 3 and 6-8. Younger (<50 years) participants generally demonstrated higher knowledge scores, reported greater familiarity with the Eatwell Guide, and more barriers across practical (e.g. time, cost, meal planning), environmental (e.g. unhealthy food availability), psychological (e.g. mood, comfort eating), and social domains (e.g. celebrations, cultural traditions). They also more strongly endorsed facilitators across educational (e.g. opportunities to learn, public health promotion), practical (e.g. budget-friendly meals, portion size examples, healthier foods at work), motivational (e.g. health, mental health, and weight management outcomes), and environmental facilitators (e.g. restaurant options, cheaper healthy food, family and friends support). Despite this, older participants (>50 years) were more likely to perceive the guidelines as personally relevant and to report at least occasional adherence.

Responses also consistently differed between participants of different ethnicities. Non-white participants scored lower on both Eatwell Guide knowledge scores, reported lower familiarity with the guidelines, and reported more barriers across knowledge-based (e.g. understanding guidelines, food labelling), practical (e.g. time, meal planning), environmental (e.g. unhealthy food availability), and cultural domains (e.g. cultural traditions, cultural food recommendations from healthcare professionals). Non-white participants also more strongly endorsed facilitators across almost all domains, particularly practical (e.g. budget meals, affordable cooking), cultural (e.g. culturally appropriate food options), and motivational facilitators (e.g. health, weight management).

Sex differences were less consistent. Females demonstrated higher Eatwell Guide knowledge, greater familiarity with the guidelines, and a greater understanding of the importance of the guidelines. Males reported more practical and knowledge-based barriers (e.g. understanding guidelines, cooking skills). Females reported greater psychological barriers (e.g. mood, comfort eating) and more strongly endorsed practical facilitators (e.g. budget meals, portion size examples).

BMI differences were the most limited. Participants with overweight or obesity reported lower perceived dietary alignment, more psychological and environmental barriers (e.g. controlling eating, mood, expense), and more strongly endorsed outcome-focused facilitators (e.g. weight management, mental health improvement, enjoyable food).

## DISCUSSION

This is the first study to characterise knowledge of UK dietary guidelines and perceived barriers and facilitators to adherence in a nationally representative UK sample. Whilst most participants reported some familiarity with UK healthy eating guidelines, this was markedly lower for the Eatwell Guide specifically, with almost half reporting no familiarity at all. Despite this, knowledge of Eatwell Guide specific recommendations was moderate under strict scoring and improved under liberal scoring, suggesting good general awareness of recommendations. Specific knowledge of the underpinning nutrient-based dietary guidelines (DRVs) was very poor but improved to show moderate awareness of these recommendations in more liberal scoring. Barriers to dietary guideline adherence were predominantly social, environmental and psychological in nature, whilst perceived facilitators were primarily economic and health outcome focused. These findings have important implications for the design of behaviour change interventions aimed at improving dietary guideline adherence in the UK population.

Knowledge of broader nutrient-based dietary guidelines (DRVs) was generally poor, although given these guidelines are not intended for direct public communication, this was perhaps unsurprising. However, knowledge of Eatwell Guide specific recommendations was moderate to high, which was unexpected given that almost half of participants reported no familiarity with the Eatwell Guide at all. This suggests that many participants possess a reasonable awareness of general healthy eating principles, despite limited familiarity with the Eatwell Guide as a specific tool. This may reflect widespread diffusion of core dietary messages (although not necessarily the Eatwell Guide ‘branding’ of these messages) across public health communications. Nutrition knowledge has been positively, albeit weakly, associated with dietary quality previously (Spronk et al., 2014) and is considered an important component of health literacy, which is itself associated with broader health outcomes (Batterham et al., 2016). However, evidence suggests that knowledge alone is insufficient for behaviour change (Jezewska-Zychowicz and Plichta, 2022). This is supported by the present study in which ∼6% of participants reported following guidelines consistently, and a third reported no intention to do so. These data highlight the importance of understanding the broader barriers and facilitators to dietary guideline adherence.

The most commonly reported barriers were social, environmental and psychological, with practical barriers such as cooking skills and finding appropriate foods among the least reported. This pattern is broadly consistent with previous UK research identifying taste preferences, daily habits, and environmental factors as key barriers to healthy eating (Mc Morrow et al., 2016; Michaelidou et al., 2012). In the context of COM-B, these findings suggest that physical capability is not the primary obstacle to dietary guideline adherence, and that barriers appear to predominantly reflect deficits in motivation and opportunity. Psychological barriers including mood, comfort eating and difficulty controlling eating align with the automatic motivation component of COM-B (Michie et al., 2011), suggesting that eating behaviour is frequently driven by habitual and emotional processes that override conscious dietary intentions. This is supported by previous research which identified difficulty changing established eating habits as among the barriers most strongly associated with lower dietary adherence in UK adults (Michaelidou et al., 2012). Environmental barriers, notably the availability of unhealthy foods and the perceived cost of healthy eating, suggest that individual behaviour change alone may be insufficient without broader changes to the food environment (Pineda et al., 2024).

The most commonly reported facilitators were economic, with cheaper healthy foods and budget meal options most strongly endorsed. Notably, whilst only 25.5% of participants identified the cost of healthy eating as a direct barrier, 74.0% endorsed cheaper healthy foods as a facilitator, suggesting that food affordability may be influencing dietary behaviour even among those who do not explicitly frame cost as a personal barrier. This discrepancy may reflect a tendency to underreport financial constraints as a barrier whilst simultaneously recognising affordability as a key structural enabler of healthier eating. Taken together, these findings reinforce affordability and access to healthy food as central structural determinants of dietary adherence, particularly pertinent in the context of evidence that healthier foods have been consistently more expensive than less healthy alternatives in the UK over the past decade, with the recent cost-of-living crisis potentially exacerbating inequalities in dietary quality (Hoenink et al., 2024). The strong endorsement of health outcome motivators (weight management, mental health and general health improvement) is consistent with evidence that health benefits, prevention of illness and wellbeing are among the most consistently identified facilitators of healthy behaviour change (Kelly et al., 2016). Notably, intrinsic health motives have been shown to be the strongest predictor of healthy eating intention and behaviour in UK adults (Michaelidou et al., 2012), suggesting that public health communications framing dietary guideline adherence in terms of tangible personal health benefits may be particularly effective in motivating behaviour change. Ease of understanding the Eatwell Guide was among the least endorsed facilitators, with fewer than a third of participants agreeing this would facilitate their adherence. This suggests the tool itself may not be well positioned to support behaviour change in its current form, consistent with evidence that people are often aware of what changes are needed but lack the specific, practical information and strategies to implement them (Kelly et al., 2016).

Whilst statistically significant differences were observed across several demographic subgroups (particularly for age), absolute differences were generally small and should be interpreted with appropriate caution. The most notable and consistent equity-related findings related to ethnicity. Non-white participants demonstrated lower knowledge of Eatwell Guide recommendations, lower familiarity with UK dietary guidelines, and reported more barriers across practical, environmental and cultural domains. This aligns with qualitative evidence from Ojo et al., (2023a), who found that ethnically diverse communities in England encountered substantial structural and cultural barriers to healthy eating and perceived UK dietary guidelines as not designed for or applicable to them. Whilst culturally adapted versions of the Eatwell Guide exist (Jay, 2021; Saint Hill, 2023), awareness of these resources was very low even among non-white participants in the present study (13.4%), suggesting that current dissemination strategies are not effectively reaching the communities these resources were designed to support. Notably, Ojo et al.(Ojo et al., 2023a) also found that individuals from minority ethnic communities were more likely to seek dietary information through social media and community or religious networks, rather than through official government channels. This may inform the development of future dissemination strategies for these populations.

Findings from the present study have several important implications for policy and practice (Figure 1). Despite GPs and healthcare professionals being the most preferred source of dietary information, only 14.3% of participants currently receive dietary guidance from this source. This represents a substantial gap between preference and provision. In line with the Make Every Contact Count approach (NHS England, 2016), the integration of brief dietary advice into routine primary care contacts may represent a low cost, feasible intervention opportunity. The strong endorsement of economic facilitators (e.g., cheaper healthy foods and budget meal plans), alongside the prominent role of environmental barriers (e.g., availability of unhealthy foods), reinforces the case for policy responses targeting food affordability and availability. Whilst socioeconomic variation in Eatwell Guide adherence has been reported previously (Griffiths et al., 2025b), the distribution of the present sample precluded meaningful subgroup analysis by SES. Nevertheless, evidence suggests that environmental and contextual barriers are disproportionately prevalent among disadvantaged UK adults (Briazu et al., 2024), and that individual behaviour change interventions are unlikely to be sufficient without complementary structural changes to the food environment. The very low awareness of culturally adapted

**Figure 1.**
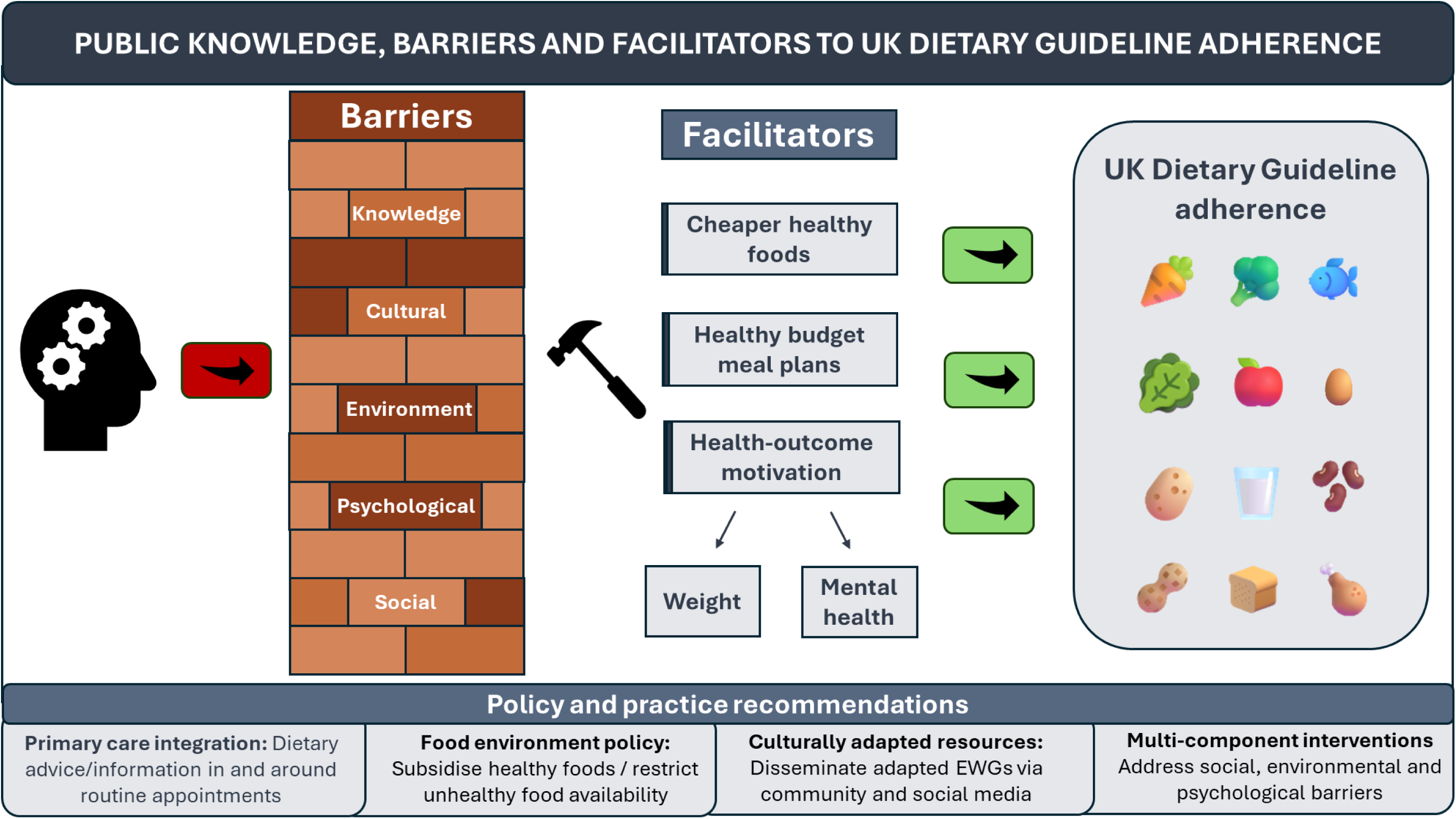
**Summary of key findings and policy and practice recommendations**

Eatwell Guides highlights a potential need to improve dissemination through community, religious and social media channels, which evidence suggests are more likely to be used by ethnic minority communities than official government sources (Ojo et al., 2023a, 2023b). However, it is acknowledged that more work may first be needed to determine the health effects of adhering to these culturally adapted versions of the Eatwell Guide. Given that psychological barriers including mood, comfort eating and difficulty controlling eating were among the most commonly endorsed, interventions should also specifically target the emotional and habitual drivers of dietary behaviour, with evidence suggesting that behaviour change techniques targeting self-regulation, values and future outcomes may be particularly effective in this regard (Power et al., 2025). Finally, given that ease of understanding the Eatwell Guide was among the least endorsed facilitators, and that barriers in the present study spanned motivational and opportunity domains of the COM-B model, interventions targeting knowledge alone are unlikely to be sufficient. Effective behaviour change strategies should address affordability and structural barriers, frame adherence in terms of tangible health outcomes, and adopt multi-component approaches spanning the motivational and opportunity domains identified in the present study (Michie et al., 2011).

To our knowledge, this is the first study to simultaneously characterise knowledge of UK nutrient and food-based dietary guidelines and perceived barriers and facilitators to adherence in a nationally representative sample. The large sample size and representation across age, sex and ethnicity strengthens generalisability. In addition, the use of both strict and liberal knowledge scoring provided a more nuanced picture of dietary knowledge than a single score would allow. However, several limitations should be considered. First, the cross-sectional design means it is not possible to determine the directionality of relationships between knowledge, barriers and adherence. Second, reliance on self-reported measures throughout introduces the possibility of social desirability bias, particularly for dietary adherence data (Kowalski et al., 2025). Third, the knowledge assessment and barrier and facilitator questions were developed specifically for this study (something we deemed necessary given a lack of current tools available for this purpose) and have not been formally validated, meaning it is not possible to confirm that items accurately capture the constructs they were designed to measure. Future research should develop and validate tools for assessing UK dietary guideline knowledge and barriers to adherence. Fourth, the limited spread of socioeconomic status in the present sample, precluded meaningful subgroup analysis by SES despite well-documented socioeconomic variation in dietary behaviour. Fifth, whilst the survey approach enabled large-scale data collection, a qualitative approach using interviews or focus groups may have provided richer insight into the lived experience of the UK population. Follow up studies using such designs are therefore warranted to complement the findings observed here, and could use our results to help shape interview templates. Finally, whilst a small number of participants in the present study reported current use of GLP-1 receptor agonist medications, the sample was insufficient to examine this group separately. This represents an important avenue for future research, given that individuals using these medications consume significantly reduced food volumes, potentially increasing nutritional inadequacy risk if food choices do not align closely with healthy eating guidelines. Evidence suggests that dietary support for GLP-1 users is currently inadequate (Griffiths et al., 2025c), highlighting an urgent need for research into how dietary guidance can be effectively tailored and delivered to this growing population.

In conclusion, this study demonstrates that whilst knowledge of Eatwell Guide recommendations is moderate to high, familiarity with the guide itself is low and knowledge of broader dietary guidelines remains poor. Barriers to adherence are predominantly social, environmental and psychological, and facilitators primarily economic and health outcome focused. Significant equity concerns were identified, particularly relating to ethnicity, with non-white participants facing disproportionate barriers. Improving dietary guideline adherence in the UK requires structural changes to the food environment, improved dissemination of culturally adapted resources through community channels, integration of dietary guidance into routine primary care, and multi-component behaviour change interventions that extend beyond knowledge alone.

## Data Availability

All data produced in the present study are available upon reasonable request to the authors

