## Supplementary materials for "PUBLIC KNOWLEDGE, BARRIERS AND FACILITATORS TO UK DIETARY GUIDELINE ADHERENCE: A NATIONALLY REPRESENTATIVE SURVEY"

**Supplementary Table 1. Response distributions for Eatwell Guide specific knowledge questions under strict and liberal scoring.**

| Question | Response | n | % | Strict % correct | Liberal % correct |
|---|---|---|---|---|---|
| **What proportion of daily food intake should come from starchy carbohydrates?** | About one quarter ✓ | 433 | 43.2 | 38.8 | 88.7 |
| | About one third ✓✓ | 389 | 38.8 | | |
| | About one half ✓ | 68 | 6.8 | | |
| | About three quarters | 18 | 1.8 | | |
| | Avoid | 31 | 3.1 | | |
| | No recommendation | 64 | 6.4 | | |
| **What type of starchy carbohydrates does the Eatwell Guide recommend?** | White/refined carbohydrates | 9 | 0.9 | 94.6 | 94.6 |
| | Wholegrain or higher fibre varieties ✓✓ | 949 | 94.6 | | |
| | Low-carbohydrate options | 28 | 2.8 | | |
| | Avoid all carbohydrates | 6 | 0.6 | | |
| | No recommendation | 11 | 1.1 | | |
| **What types of protein does the Eatwell Guide recommend?** | Only animal sources | 16 | 1.6 | 90.8 | 90.8 |
| | Variety including beans, pulses, fish, eggs, lean meat ✓✓ | 911 | 90.8 | | |
| | Only plant-based sources | 16 | 1.6 | | |
| | No recommendation | 60 | 6.0 | | |
| **What does the Eatwell Guide recommend about oils and spreads?** | Avoid | 49 | 4.9 | 70.4 | 70.4 |
| | Saturated fats like butter and coconut oil | 162 | 16.2 | | |
| | Unsaturated oils and spreads in small amounts ✓✓ | 706 | 70.4 | | |
| | As much as needed for cooking and flavour | 8 | 0.8 | | |
| | No recommendation | 78 | 7.8 | | |
| **What does the Eatwell Guide recommend about dairy?** | Avoid | 19 | 1.9 | 55.4 | 55.4 |
| | Full-fat only in moderate quantities | 130 | 13.0 | | |
| | Lower-fat and lower-sugar in moderate quantities ✓✓ | 556 | 55.4 | | |
| | Full-fat only in high quantities | 7 | 0.7 | | |
| | Lower-fat and lower-sugar in high quantities | 57 | 5.7 | | |
| | No recommendation | 234 | 23.3 | | |
| **How much should you consume foods high in fat, salt and sugar (e.g. cakes, crisps, sweets)?** | High | 3 | 0.3 | 53.9 | 93.7 |
| | Moderate | 23 | 2.3 | | |
| | Low ✓✓ | 595 | 59.3 | | |
| | Avoid ✓ | 345 | 34.4 | | |
| | No recommendation | 37 | 3.7 | | |
| **How many portions of fruit and vegetables should you consume per day?** | 2 portions | 40 | 4.0 | 84.8 | 90.4 |
| | 3 portions | 50 | 5.0 | | |
| | 4 portions ✓ | 27 | 2.7 | | |
| | 5 portions ✓✓ | 851 | 84.8 | | |
| | 6 portions ✓ | 29 | 2.9 | | |
| | No recommendation | 6 | 0.6 | | |
| | Consume as much as you like | 19 | 1.9 | 18.9 | 47.3 |

| | | | | | |
|---|---|---|---|---|---|
| **What is the maximum recommended daily intake of fruit juice or smoothies?** | Up to 100ml per day ✓ | 154 | 15.4 | | |
| | Up to 150ml per day ✓✓ | 190 | 18.9 | | |
| | Up to 200ml per day ✓ | 130 | 13.0 | | |
| | Up to 250ml per day | 111 | 11.1 | | |
| | Should be avoided completely | 84 | 8.4 | | |
| | No recommendation | 315 | 31.4 | | |
| **How many portions of fish per week does the Eatwell Guide recommend?** | 1 portion ✓ | 87 | 8.7 | 47.3 | 75.5 |
| | 2 portions ✓✓ | 474 | 47.3 | | |
| | 3 portions ✓ | 196 | 19.5 | | |
| | 4 portions | 41 | 4.1 | | |
| | No recommendation | 205 | 20.4 | | |
| **How many portions of oily fish per week does the Eatwell Guide recommend?** | 0 portions | 36 | 3.6 | 40.0 | 73.6 |
| | At least 1 portion ✓✓ | 401 | 40.0 | | |
| | At least 2 portions ✓ | 337 | 33.6 | | |
| | At least 3 portions | 90 | 9.0 | | |
| | No recommendation | 139 | 13.9 | | |
| **What is the recommended daily calorie intake for an adult male?** | 3000kcal ✓ | 90 | 9.0 | 67.5 | 96 |
| | 2500kcal ✓✓ | 677 | 67.5 | | |
| | 2000kcal ✓ | 194 | 19.3 | | |
| | 1500kcal | 25 | 2.5 | | |
| | 1000kcal | 3 | 0.3 | | |
| | No recommendation | 14 | 1.4 | | |
| **What is the recommended daily calorie intake for an adult female?** | 3000kcal | 8 | 0.8 | 68.9 | 95.6 |
| | 2500kcal ✓ | 59 | 5.9 | | |
| | 2000kcal ✓✓ | 691 | 68.9 | | |
| | 1500kcal ✓ | 209 | 20.8 | | |
| | 1000kcal | 20 | 2.0 | | |
| | No recommendation | 16 | 1.6 | | |
| **Which of the following count towards your 5-a-day? (select all that apply)** | *Items that count: fresh, tinned, and frozen fruit and vegetables; beans and pulses; dried fruit; fruit juice (up to 150ml); smoothies. Items that do not count: potatoes Strict: all correct items selected, no incorrect items Liberal: all correctly selected or only one error (missing or incorrectly selected)* | | | 7.1 | 27.1 |
| **Which of the following count towards your recommended fluid intake? (select all that apply)** | *Items that count: water, tea, coffee, low-sugar squash, milk, herbal tea, fruit juice (up to 150ml) Items that do not count: alcoholic drinks, sugary drinks, energy drinks, fruit squash Strict: all correct items selected, no incorrect items Liberal: all correct or only one error* | | | 2.2 | 15.8 |

*Note. n = 1003. ✓✓ = the correct answer and awarded a point in strict scoring. ✓ = awarded a point under liberal scoring only. Strict scoring awards 1 point for the correct answer only. Liberal scoring awards 1 point for the correct answer or adjacent/near-correct responses. For the fruit and vegetable count and fluid types questions, strict scoring requires all correct items selected (and no incorrect items); liberal scoring awards 1 point if all correct or only one error. As n and % are identical for strict and liberal scoring for most items (the response options are the same; only the scoring differs), the columns show how many participants selected each response under each scoring approach.*

**Supplementary Table 2. Response distributions for broader dietary guideline knowledge questions under strict and liberal scoring.**

| Question | Response | n | % | Strict % correct | Liberal % correct |
|---|---|---|---|---|---|
| **What percentage of daily calories should come from carbohydrates?** | 20% | 298 | 29.7 | **16.3** | **65.9** |
| | 40% ✓ | 432 | 43.1 | | |
| | 50% ✓✓ | 163 | 16.3 | | |
| | 60% ✓ | 66 | 6.6 | | |
| | 80% | 6 | 0.6 | | |
| | No recommendation | 38 | 3.8 | | |
| **What percentage of daily calories should come from free sugars?** | ≤6% ✓ | 132 | 13.2 | **25.7** | **57.6** |
| | ≤5% ✓✓ | 258 | 25.7 | | |
| | <4% ✓ | 188 | 18.7 | | |
| | <2% | 325 | 32.4 | | |
| | 0% | 37 | 3.7 | | |
| | No recommendation | 63 | 6.3 | | |
| **How many grams of fibre should you consume per day?** | 15g | 57 | 5.7 | **32.2** | **73.7** |
| | 20g | 177 | 17.6 | | |
| | 25g ✓ | 313 | 31.2 | | |
| | 30g ✓✓ | 323 | 32.2 | | |
| | 35g ✓ | 103 | 10.3 | | |
| | No recommendation | 30 | 3.0 | | |
| **What percentage of daily calories should come from protein?** | 10% ✓ | 42 | 4.2 | **12.1** | **49.1** |
| | 15% ✓✓ | 121 | 12.1 | | |
| | 20% ✓ | 329 | 32.8 | | |
| | 30% | 303 | 30.2 | | |
| | 35% | 175 | 17.4 | | |
| | No recommendation | 33 | 3.3 | | |
| **What percentage of daily calories should come from total fat?** | 25% or less | 718 | 71.6 | **4.5** | **22.8** |
| | 30% or less ✓ | 177 | 17.6 | | |
| | 35% or less ✓✓ | 45 | 4.5 | | |
| | 40% or less ✓ | 7 | 0.7 | | |
| | 45% or less | 4 | 0.4 | | |
| | No recommendation | 52 | 5.2 | | |
| **What percentage of daily calories should come from saturated fat?** | ≤9% ✓ | 720 | 71.8 | **11.3** | **86.1** |
| | ≤11% ✓✓ | 113 | 11.3 | | |
| | ≤13% ✓ | 31 | 3.1 | | |
| | ≤15% | 22 | 2.2 | | |
| | ≤17% | 3 | 0.3 | | |
| | No recommendation | 114 | 11.4 | | |
| **What is the maximum recommended daily salt intake?** | 4g | 466 | 46.5 | **18.2** | **47.2** |
| | 5g ✓ | 262 | 26.1 | | |
| | 6g ✓✓ | 183 | 18.2 | | |
| | 7g ✓ | 28 | 2.8 | | |
| | 8g | 18 | 1.8 | | |
| | No recommendation | 46 | 4.6 | | |

| | | | | | |
|---|---|---|---|---|---|
| **What is the maximum recommended daily intake of red and processed meat?** | 50g ✓ | 292 | 29.1 | **23.3** | **68.8** |
| | 70g ✓✓ | 234 | 23.3 | | |
| | 90g ✓ | 164 | 16.4 | | |
| | 120g | 97 | 9.7 | | |
| | 140g | 19 | 1.9 | | |
| | No recommendation | 197 | 19.6 | | |

*Note. n = 1003. ✓✓ = the correct answer and awarded a point in strict scoring. ✓ = awarded a point under liberal scoring only. Strict scoring awards 1 point for the correct answer only. Liberal scoring awards 1 point for the correct answer or adjacent/near-correct responses*

**Supplementary Table 3. Response distributions and group comparisons for perceptions of UK Government healthy eating guidelines.**

| Question | Response | Overall (n=1003) | | Age | | | | | Sex | | | | | Ethnicity | | | | | BMI | | | | |
|---|---|---|---|---|---|---|---|---|---|---|---|---|---|---|---|---|---|---|---|---|---|---|---|
| | | | | Under 50 | | 50 and over | | p | Male | | Female | | p | White | | Non-white | | p | Healthy | | Overweight/ obesity | | p |
| | | n | % | n | % | n | % | | n | % | n | % | | n | % | n | % | | n | % | n | % | |
| **How familiar are you with UK Government healthy eating guidelines?** | Not at all familiar | 240 | 23.9 | 127 | 23.4 | 113 | 24.5 | 0.216 | 138 | 28.6 | 101 | 19.6 | **0.002** | 187 | 21.9 | 51 | 35.9 | **0.006** | 104 | 24.8 | 127 | 23.1 | 0.928 |
| | Somewhat familiar | 654 | 65.2 | 347 | 64.0 | 307 | 66.6 | | 298 | 61.7 | 352 | 68.3 | | 576 | 67.4 | 75 | 52.8 | | 266 | 63.3 | 365 | 66.5 | |
| | Highly familiar | 89 | 8.9 | 54 | 10.0 | 35 | 7.6 | | 38 | 7.9 | 51 | 9.9 | | 74 | 8.7 | 14 | 9.9 | | 41 | 9.8 | 47 | 8.6 | |
| | Extremely familiar | 20 | 2.0 | 14 | 2.6 | 6 | 1.3 | | 9 | 1.9 | 11 | 2.1 | | 18 | 2.1 | 2 | 1.4 | | 9 | 2.1 | 10 | 1.8 | |
| **How familiar are you with the Eatwell Guide?** | Not at all familiar | 494 | 49.3 | 230 | 42.4 | 264 | 57.3 | **<0.001** | 274 | 56.7 | 217 | 42.1 | **<0.001** | 418 | 48.9 | 73 | 51.4 | 0.714 | 200 | 47.6 | 273 | 49.7 | 0.223 |
| | Somewhat familiar | 396 | 39.5 | 232 | 42.8 | 164 | 35.6 | | 168 | 34.8 | 226 | 43.9 | | 342 | 40.0 | 52 | 36.6 | | 162 | 38.6 | 225 | 41.0 | |
| | Highly familiar | 77 | 7.7 | 54 | 10.0 | 23 | 5.0 | | 31 | 6.4 | 46 | 8.9 | | 66 | 7.7 | 11 | 7.7 | | 41 | 9.8 | 34 | 6.2 | |
| | Extremely familiar | 36 | 3.6 | 26 | 4.8 | 10 | 2.2 | | 10 | 2.1 | 26 | 5.0 | | 29 | 3.4 | 6 | 4.2 | | 17 | 4.0 | 17 | 3.1 | |
| **Are you aware of alternative EWG versions which account for cultural differences?** | Yes | 57 | 5.7 | 40 | 7.4 | 17 | 3.7 | **0.017** | 29 | 6.0 | 28 | 5.4 | 0.803 | 37 | 4.3 | 19 | 13.4 | **<0.001** | 26 | 6.2 | 29 | 5.3 | 0.642 |
| | No | 946 | 94.3 | 502 | 92.6 | 444 | 96.3 | | 454 | 94.0 | 487 | 94.6 | | 818 | 95.7 | 123 | 86.6 | | 394 | 93.8 | 520 | 94.7 | |
| **Are you aware of alternative EWG versions which account for dietary preferences?** | Yes | 71 | 7.1 | 51 | 9.4 | 20 | 4.3 | **0.003** | 29 | 6.0 | 42 | 8.2 | 0.231 | 55 | 6.4 | 15 | 10.6 | 0.108 | 33 | 7.9 | 33 | 6.0 | **0.316** |
| | No | 932 | 92.9 | 491 | 90.6 | 441 | 95.7 | | 454 | 94.0 | 473 | 91.8 | | 800 | 93.6 | 127 | 89.4 | | 387 | 92.1 | 516 | 94.0 | |
| **How well does your diet align with UK Government healthy eating guidelines?** | Not at all | 60 | 6.7 | 34 | 7.0 | 26 | 6.3 | **0.049** | 28 | 6.6 | 31 | 6.6 | 0.257 | 48 | 6.2 | 12 | 10.1 | 0.673 | 13 | 3.5 | 46 | 9.2 | **<0.001** |
| | Slightly | 277 | 30.8 | 164 | 33.5 | 113 | 27.6 | | 139 | 32.6 | 136 | 29.0 | | 241 | 31.0 | 34 | 28.6 | | 90 | 24.3 | 180 | 35.9 | |
| | Moderately | 524 | 58.4 | 273 | 55.8 | 252 | 61.5 | | 245 | 57.4 | 280 | 59.7 | | 457 | 58.8 | 67 | 56.3 | | 244 | 65.9 | 264 | 52.6 | |
| | Completely | 38 | 4.2 | 18 | 3.7 | 19 | 4.6 | | 15 | 3.5 | 22 | 4.7 | | 31 | 4.0 | 6 | 5.0 | | 23 | 6.2 | 12 | 2.4 | |
| **How important is it to adhere to UK Government healthy eating guidelines?** | Not at all important | 68 | 7.2 | 39 | 7.5 | 29 | 6.9 | 0.107 | 34 | 7.6 | 33 | 6.7 | **0.016** | 60 | 7.5 | 8 | 6.0 | 0.659 | 28 | 7.0 | 37 | 7.2 | 0.567 |
| | Slightly important | 217 | 23.0 | 131 | 25.2 | 86 | 20.3 | | 114 | 25.4 | 103 | 20.9 | | 181 | 22.5 | 35 | 26.1 | | 96 | 24.0 | 112 | 21.8 | |
| | Moderately important | 464 | 49.2 | 248 | 47.7 | 216 | 51.1 | | 222 | 49.6 | 240 | 48.8 | | 404 | 50.2 | 58 | 43.3 | | 195 | 48.8 | 255 | 49.7 | |
| | Very important | 194 | 20.6 | 102 | 19.6 | 92 | 21.7 | | 78 | 17.4 | 116 | 23.6 | | 160 | 19.9 | 33 | 24.6 | | 81 | 20.2 | 109 | 21.2 | |

| | | | | | | | | | | | | | | | | | | | | | | |
|---|---|---|---|---|---|---|---|---|---|---|---|---|---|---|---|---|---|---|---|---|---|---|
| **How relevant are UK Government healthy eating guidelines to you personally?** | Not at all relevant | 78 | 8.7 | 40 | 8.2 | 38 | 9.2 | **0.014** | 36 | 8.3 | 39 | 8.5 | 0.146 | 67 | 8.7 | 10 | 8.5 | 0.985 | 31 | 8.1 | 40 | 8.2 | 0.066 |
| | Slightly relevant | 237 | 26.4 | 144 | 29.6 | 93 | 22.6 | | 121 | 27.8 | 116 | 25.3 | | 202 | 26.1 | 32 | 27.1 | | 115 | 30.1 | 116 | 23.9 | |
| | Moderately relevant | 444 | 49.5 | 243 | 50.0 | 201 | 48.9 | | 223 | 51.3 | 220 | 48.0 | | 386 | 49.9 | 57 | 48.3 | | 183 | 47.9 | 247 | 50.8 | |
| | Completely relevant | 138 | 15.4 | 59 | 12.1 | 79 | 19.2 | | 55 | 12.6 | 83 | 18.1 | | 119 | 15.4 | 19 | 16.1 | | 53 | 13.9 | 83 | 17.1 | |
| **Do you follow UK Government healthy eating guidelines?** | Yes, consistently | 56 | 5.6 | 23 | 4.8 | 26 | 6.5 | **<0.001** | 19 | 4.6 | 30 | 6.5 | 0.632 | 43 | 5.7 | 5 | 4.1 | **0.007** | 30 | 8.1 | 17 | 3.5 | **0.031** |
| | Yes, occasionally | 408 | 40.7 | 179 | 37.1 | 179 | 45.0 | | 167 | 40.4 | 191 | 41.3 | | 313 | 41.5 | 44 | 36.4 | | 148 | 40.1 | 202 | 42.2 | |
| | No, but I intend to | 205 | 20.5 | 127 | 26.3 | 53 | 13.3 | | 86 | 20.8 | 93 | 20.1 | | 140 | 18.5 | 39 | 32.2 | | 70 | 19.0 | 104 | 21.7 | |
| | No, and I don't intend to | 334 | 33.3 | 153 | 31.7 | 140 | 35.2 | | 141 | 34.1 | 149 | 32.2 | | 259 | 34.3 | 33 | 27.3 | | 121 | 32.8 | 156 | 32.6 | |

Note. n = 1003. Age: Under 50 (n=542) vs 50 and over (n=461). Sex: Male (n=483) vs Female (n=515). Ethnicity: White (n=855) vs Non-white (n=142). BMI: Healthy weight (n=420) vs Overweight/obesity (n=549). Not sure responses excluded from ordinal analyses but reported in overall column. Bold p values indicate statistical significance (p<0.05). p values from Mann-Whitney U test (ordinal variables) or Chi-square test (nominal variables)

**Supplementary Table 4. Current sources of information about UK Government healthy eating guidelines, ranked by frequency.**

| Source of information | n | % |
|---|---|---|
| No information from any source | 452 | 45.1 |
| Government websites (e.g. NHS, GOV.UK) | 193 | 19.2 |
| School or education | 184 | 18.3 |
| GP or other healthcare professional | 143 | 14.3 |
| Social media | 122 | 12.2 |
| Supermarkets | 110 | 11.0 |
| TV or radio | 108 | 10.8 |
| Friends or family | 85 | 8.5 |
| Not sure | 84 | 8.4 |
| Other websites | 45 | 4.5 |
| Workplace | 32 | 3.2 |
| Diet clubs or slimming groups | 30 | 3.0 |
| Weight loss prescriber | 14 | 1.4 |
| Other | 6 | 0.6 |

*Note. n = 1003. Participants were asked to select all sources from which they had received information about UK Government healthy eating guidelines. Percentages do not sum to 100% as participants could select multiple sources*

**Supplementary Table 5. Preferred sources of information about UK Government healthy eating guidelines, ranked by mean preference score.**

| Source of information | Mean | SD |
|---|---|---|
| GP or other healthcare professional | 3.07 | 2.18 |
| Government websites (e.g. NHS, GOV.UK) | 3.51 | 2.38 |
| Supermarkets | 4.39 | 2.50 |
| TV or radio | 5.47 | 2.54 |
| School or education | 5.51 | 2.83 |
| Friends or family | 5.69 | 2.40 |
| Social media | 5.92 | 3.09 |
| Newspapers or magazines | 6.49 | 2.37 |
| Workplace | 7.37 | 2.20 |
| Diet clubs or slimming groups | 7.58 | 2.37 |

*Note. n = 1003. Participants ranked 10 sources from 1 (most preferred) to 10 (least preferred). Lower mean scores indicate greater preferences*

**Supplementary Table 6. Response distributions and group comparisons for perceived barriers to following UK Government healthy eating guidelines.**

| Question | Response | Overall (n=1003) | | Age | | | | | Sex | | | | | Ethnicity | | | | | BMI | | | | |
|---|---|---|---|---|---|---|---|---|---|---|---|---|---|---|---|---|---|---|---|---|---|---|---|
| | | n | % | Under 50 | | 50 and over | | p | Male | | Female | | p | White | | Non-white | | p | Healthy weight | | Overweight /obesity | | p |
| | | | | n | % | n | % | | n | % | n | % | | n | % | n | % | | n | % | n | % | |
| **I find it difficult to understand the UK Government healthy eating guidelines** | Strongly agree | 34 | 3.4 | 16 | 3.0 | 18 | 3.9 | 0.856 | 25 | 5.2 | 9 | 1.7 | **0.001** | 27 | 3.2 | 7 | 4.9 | **<0.001** | 9 | 2.1 | 25 | 4.6 | 0.131 |
| | Agree | 158 | 15.8 | 90 | 16.6 | 68 | 14.8 | | 84 | 17.4 | 71 | 13.8 | | 113 | 13.2 | 43 | 30.3 | | 61 | 14.5 | 89 | 16.2 | |
| | Neither agree nor disagree | 275 | 27.4 | 142 | 26.2 | 133 | 28.9 | | 138 | 28.6 | 136 | 26.4 | | 239 | 28.0 | 34 | 23.9 | | 120 | 28.6 | 144 | 26.2 | |
| | Disagree | 449 | 44.8 | 249 | 45.9 | 200 | 43.4 | | 201 | 41.6 | 247 | 48.0 | | 401 | 46.9 | 46 | 32.4 | | 185 | 44.0 | 251 | 45.7 | |
| | Strongly disagree | 87 | 8.7 | 45 | 8.3 | 42 | 9.1 | | 35 | 7.2 | 52 | 10.1 | | 75 | 8.8 | 12 | 8.5 | | 45 | 10.7 | 40 | 7.3 | |
| **I find it difficult to choose the right foods to meet the guidelines** | Strongly agree | 15 | 1.5 | 11 | 2.0 | 4 | 0.9 | 0.391 | 9 | 1.9 | 5 | 1.0 | **<0.001** | 12 | 1.4 | 3 | 2.1 | **0.017** | 7 | 1.7 | 7 | 1.3 | 0.585 |
| | Agree | 108 | 10.8 | 61 | 11.3 | 47 | 10.2 | | 60 | 12.4 | 47 | 9.1 | | 82 | 9.6 | 26 | 18.3 | | 45 | 10.7 | 57 | 10.4 | |
| | Neither agree nor disagree | 182 | 18.1 | 96 | 17.7 | 86 | 18.7 | | 105 | 21.7 | 76 | 14.8 | | 148 | 17.3 | 31 | 21.8 | | 71 | 16.9 | 102 | 18.6 | |
| | Disagree | 531 | 52.9 | 288 | 53.1 | 243 | 52.7 | | 240 | 49.7 | 290 | 56.3 | | 472 | 55.2 | 56 | 39.4 | | 221 | 52.6 | 295 | 53.7 | |
| | Strongly disagree | 167 | 16.7 | 86 | 15.9 | 81 | 17.6 | | 69 | 14.3 | 97 | 18.8 | | 141 | 16.5 | 26 | 18.3 | | 76 | 18.1 | 88 | 16.0 | |
| **I find it difficult to know what counts as the right portion sizes** | Strongly agree | 39 | 3.9 | 30 | 5.5 | 9 | 2.0 | **0.005** | 18 | 3.7 | 20 | 3.9 | 0.870 | 28 | 3.3 | 11 | 7.7 | **<0.001** | 13 | 3.1 | 24 | 4.4 | 0.206 |
| | Agree | 202 | 20.1 | 123 | 22.7 | 79 | 17.1 | | 97 | 20.1 | 102 | 19.8 | | 152 | 17.8 | 49 | 34.5 | | 85 | 20.2 | 103 | 18.8 | |
| | Neither agree nor disagree | 165 | 16.5 | 86 | 15.9 | 79 | 17.1 | | 80 | 16.6 | 85 | 16.5 | | 142 | 16.6 | 21 | 14.8 | | 63 | 15.0 | 99 | 18.0 | |
| | Disagree | 463 | 46.2 | 233 | 43.0 | 230 | 49.9 | | 225 | 46.6 | 237 | 46.0 | | 410 | 48.0 | 50 | 35.2 | | 193 | 46.0 | 259 | 47.2 | |
| | Strongly disagree | 134 | 13.4 | 70 | 12.9 | 64 | 13.9 | | 63 | 13.0 | 71 | 13.8 | | 123 | 14.4 | 11 | 7.7 | | 66 | 15.7 | 64 | 11.7 | |
| **I don't have the cooking skills needed to follow the guidelines** | Strongly agree | 36 | 3.6 | 23 | 4.2 | 13 | 2.8 | **0.004** | 24 | 5.0 | 11 | 2.1 | **0.002** | 28 | 3.3 | 8 | 5.6 | **0.002** | 14 | 3.3 | 18 | 3.3 | 0.166 |
| | Agree | 87 | 8.7 | 61 | 11.3 | 26 | 5.6 | | 45 | 9.3 | 41 | 8.0 | | 71 | 8.3 | 14 | 9.9 | | 28 | 6.7 | 56 | 10.2 | |
| | Neither agree nor disagree | 89 | 8.9 | 45 | 8.3 | 44 | 9.5 | | 53 | 11.0 | 36 | 7.0 | | 67 | 7.8 | 20 | 14.1 | | 39 | 9.3 | 44 | 8.0 | |
| | Disagree | 406 | 40.5 | 223 | 41.1 | 183 | 39.7 | | 193 | 40.0 | 212 | 41.2 | | 345 | 40.4 | 60 | 42.3 | | 167 | 39.8 | 228 | 41.5 | |
| | Strongly disagree | 385 | 38.4 | 190 | 35.1 | 195 | 42.3 | | 168 | 34.8 | 215 | 41.7 | | 344 | 40.2 | 40 | 28.2 | | 172 | 41.0 | 203 | 37.0 | |
| **I don't have the cooking tools or equipment needed to follow the guidelines** | Strongly agree | 13 | 1.3 | 11 | 2.0 | 2 | 0.4 | **0.002** | 7 | 1.4 | 6 | 1.2 | **0.031** | 11 | 1.3 | 2 | 1.4 | **0.048** | 6 | 1.4 | 6 | 1.1 | 0.758 |
| | Agree | 46 | 4.6 | 32 | 5.9 | 14 | 3.0 | | 25 | 5.2 | 21 | 4.1 | | 36 | 4.2 | 10 | 7.0 | | 18 | 4.3 | 27 | 4.9 | |
| | Neither agree nor disagree | 35 | 3.5 | 20 | 3.7 | 15 | 3.3 | | 19 | 3.9 | 16 | 3.1 | | 29 | 3.4 | 5 | 3.5 | | 16 | 3.8 | 17 | 3.1 | |
| | Disagree | 406 | 40.5 | 228 | 42.1 | 178 | 38.6 | | 206 | 42.7 | 197 | 38.3 | | 339 | 39.6 | 64 | 45.1 | | 169 | 40.2 | 218 | 39.7 | |
| | Strongly disagree | 503 | 50.1 | 251 | 46.3 | 252 | 54.7 | | 226 | 46.8 | 275 | 53.4 | | 440 | 51.5 | 61 | 43.0 | | 211 | 50.2 | 281 | 51.2 | |
| **I find it difficult to plan meals that** | Strongly agree | 28 | 2.8 | 15 | 2.8 | 13 | 2.8 | **0.001** | 11 | 2.3 | 15 | 2.9 | **0.012** | 20 | 2.3 | 7 | 4.9 | **0.004** | 8 | 1.9 | 17 | 3.1 | **0.029** |
| | Agree | 136 | 13.6 | 94 | 17.3 | 42 | 9.1 | | 72 | 14.9 | 64 | 12.4 | | 110 | 12.9 | 24 | 16.9 | | 49 | 11.7 | 81 | 14.8 | |

| | | | | | | | | | | | | | | | | | | | | | | | |
|---|---|---|---|---|---|---|---|---|---|---|---|---|---|---|---|---|---|---|---|---|---|---|---|
| **meet the guidelines** | Neither agree nor disagree | 134 | 13.4 | 69 | 12.7 | 65 | 14.1 | | 72 | 14.9 | 61 | 11.8 | | 109 | 12.7 | 24 | 16.9 | | 43 | 10.2 | 84 | 15.3 | |
| | Disagree | 427 | 42.6 | 234 | 43.2 | 195 | 42.3 | | 214 | 44.3 | 213 | 41.4 | | 370 | 43.3 | 58 | 40.8 | | 199 | 47.4 | 218 | 39.7 | |
| | Strongly disagree | 278 | 27.7 | 130 | 24.0 | 146 | 31.7 | | 114 | 23.6 | 162 | 31.5 | | 246 | 28.8 | 29 | 20.4 | | 121 | 28.8 | 149 | 27.1 | |
| **Eating healthily in line with the guidelines is too expensive** | Strongly agree | 71 | 7.1 | 49 | 9.0 | 22 | 4.8 | **<0.001** | 29 | 6.0 | 40 | 7.8 | 0.968 | 61 | 7.1 | 10 | 7.0 | 0.349 | 29 | 6.9 | 41 | 7.5 | **0.005** |
| | Agree | 184 | 18.3 | 117 | 21.6 | 68 | 14.8 | | 90 | 18.6 | 94 | 18.3 | | 155 | 18.1 | 28 | 19.7 | | 64 | 15.2 | 115 | 20.9 | |
| | Neither agree nor disagree | 252 | 25.1 | 134 | 24.7 | 118 | 25.6 | | 126 | 26.1 | 124 | 24.1 | | 211 | 24.7 | 39 | 27.5 | | 103 | 24.5 | 140 | 25.5 | |
| | Disagree | 349 | 34.8 | 176 | 32.5 | 173 | 37.5 | | 170 | 35.2 | 179 | 34.8 | | 299 | 35.0 | 48 | 33.8 | | 148 | 35.2 | 186 | 33.9 | |
| | Strongly disagree | 147 | 14.7 | 66 | 12.2 | 80 | 17.4 | | 68 | 14.1 | 78 | 15.1 | | 129 | 15.1 | 17 | 12.0 | | 76 | 18.1 | 67 | 12.2 | |
| **I don't have enough time to eat healthily in line with the guidelines** | Strongly agree | 49 | 4.9 | 44 | 8.1 | 5 | 1.1 | **<0.001** | 24 | 5.0 | 23 | 4.5 | 0.589 | 38 | 4.4 | 11 | 7.7 | **<0.001** | 21 | 5.0 | 26 | 4.7 | 0.316 |
| | Agree | 154 | 15.4 | 120 | 22.1 | 34 | 7.4 | | 73 | 15.1 | 78 | 15.1 | | 118 | 13.8 | 34 | 23.9 | | 57 | 13.6 | 91 | 16.6 | |
| | Neither agree nor disagree | 178 | 17.7 | 106 | 19.6 | 72 | 15.6 | | 94 | 19.5 | 84 | 16.3 | | 148 | 17.3 | 27 | 19.0 | | 74 | 17.6 | 98 | 17.9 | |
| | Disagree | 407 | 40.6 | 181 | 33.4 | 226 | 49.0 | | 186 | 38.5 | 221 | 42.9 | | 356 | 41.6 | 50 | 35.2 | | 174 | 41.4 | 220 | 40.1 | |
| | Strongly disagree | 215 | 21.4 | 91 | 16.8 | 124 | 26.9 | | 106 | 21.9 | 109 | 21.2 | | 195 | 22.8 | 20 | 14.1 | | 94 | 22.4 | 114 | 20.8 | |
| **I find it difficult to find the foods I need in the shops** | Strongly agree | 20 | 2.0 | 13 | 2.4 | 7 | 1.5 | **<0.001** | 8 | 1.7 | 12 | 2.3 | **0.010** | 17 | 2.0 | 3 | 2.1 | 0.215 | 9 | 2.1 | 10 | 1.8 | 0.516 |
| | Agree | 68 | 6.8 | 54 | 10.0 | 14 | 3.0 | | 39 | 8.1 | 26 | 5.0 | | 50 | 5.8 | 16 | 11.3 | | 20 | 4.8 | 43 | 7.8 | |
| | Neither agree nor disagree | 106 | 10.6 | 59 | 10.9 | 47 | 10.2 | | 61 | 12.6 | 45 | 8.7 | | 89 | 10.4 | 15 | 10.6 | | 47 | 11.2 | 55 | 10.0 | |
| | Disagree | 466 | 46.5 | 248 | 45.8 | 218 | 47.3 | | 224 | 46.4 | 240 | 46.6 | | 403 | 47.1 | 62 | 43.7 | | 197 | 46.9 | 255 | 46.4 | |
| | Strongly disagree | 343 | 34.2 | 168 | 31.0 | 175 | 38.0 | | 151 | 31.3 | 192 | 37.3 | | 296 | 34.6 | 46 | 32.4 | | 147 | 35.0 | 186 | 33.9 | |
| **I find food labels difficult to understand** | Strongly agree | 28 | 2.8 | 15 | 2.8 | 13 | 2.8 | 0.472 | 11 | 2.3 | 17 | 3.3 | 0.547 | 24 | 2.8 | 4 | 2.8 | **0.042** | 14 | 3.3 | 13 | 2.4 | 0.409 |
| | Agree | 156 | 15.6 | 88 | 16.2 | 70 | 15.2 | | 77 | 15.9 | 79 | 15.3 | | 128 | 15.0 | 29 | 20.4 | | 64 | 15.2 | 86 | 15.7 | |
| | Neither agree nor disagree | 153 | 15.3 | 73 | 13.5 | 80 | 17.4 | | 74 | 15.3 | 79 | 15.3 | | 126 | 14.7 | 26 | 18.3 | | 54 | 12.9 | 94 | 17.1 | |
| | Disagree | 431 | 43.0 | 237 | 43.7 | 196 | 42.5 | | 204 | 42.2 | 227 | 44.1 | | 374 | 43.7 | 56 | 39.4 | | 187 | 44.5 | 233 | 42.4 | |
| | Strongly disagree | 235 | 23.4 | 129 | 23.8 | 102 | 22.1 | | 117 | 24.2 | 113 | 21.9 | | 203 | 23.7 | 27 | 19.0 | | 101 | 24.0 | 123 | 22.4 | |
| **Unhealthy food and drink is too readily available** | Strongly agree | 173 | 17.2 | 125 | 23.1 | 48 | 10.4 | **<0.001** | 83 | 17.2 | 87 | 16.9 | **0.008** | 142 | 16.6 | 29 | 20.4 | **0.016** | 72 | 17.1 | 99 | 18.0 | **0.033** |
| | Agree | 395 | 39.4 | 237 | 43.7 | 158 | 34.3 | | 212 | 43.9 | 181 | 35.1 | | 327 | 38.2 | 65 | 45.8 | | 152 | 36.2 | 225 | 41.0 | |
| | Neither agree nor disagree | 176 | 17.5 | 71 | 13.1 | 105 | 22.8 | | 85 | 17.6 | 91 | 17.7 | | 153 | 17.9 | 22 | 15.5 | | 71 | 16.9 | 101 | 18.4 | |
| | Disagree | 177 | 17.6 | 74 | 13.7 | 103 | 22.3 | | 71 | 14.7 | 106 | 20.6 | | 161 | 18.8 | 16 | 11.3 | | 78 | 18.6 | 93 | 16.9 | |
| | Strongly disagree | 82 | 8.2 | 35 | 6.5 | 47 | 10.2 | | 32 | 6.6 | 50 | 9.7 | | 72 | 8.4 | 10 | 7.0 | | 47 | 11.2 | 31 | 5.6 | |
| **Healthy food delivery services** | Strongly agree | 63 | 6.3 | 42 | 7.7 | 21 | 4.6 | **<0.001** | 33 | 6.8 | 30 | 5.8 | 0.633 | 48 | 5.6 | 14 | 9.9 | **<0.001** | 17 | 4.0 | 43 | 7.8 | 0.061 |
| | Agree | 178 | 17.7 | 133 | 24.5 | 46 | 10.0 | | 93 | 19.3 | 85 | 16.5 | | 141 | 16.5 | 37 | 26.1 | | 82 | 19.5 | 91 | 16.6 | |

| | | | | | | | | | | | | | | | | | | | | | | | |
|---|---|---|---|---|---|---|---|---|---|---|---|---|---|---|---|---|---|---|---|---|---|---|---|
| **are difficult to access** | Neither agree nor disagree | 161 | 16.1 | 78 | 14.4 | 83 | 18.0 | | 74 | 15.3 | 86 | 16.7 | | 138 | 16.1 | 21 | 14.8 | | 65 | 15.5 | 94 | 17.1 | |
| | Disagree | 298 | 29.7 | 156 | 28.8 | 142 | 30.8 | | 134 | 27.7 | 163 | 31.7 | | 254 | 29.7 | 43 | 30.3 | | 113 | 26.9 | 175 | 31.9 | |
| | Strongly disagree | 303 | 30.2 | 133 | 24.5 | 169 | 36.7 | | 149 | 30.8 | 151 | 29.3 | | 274 | 32.0 | 27 | 19.0 | | 143 | 34.0 | 146 | 26.6 | |
| **Social occasions and celebrations make it difficult to follow the guidelines** | Strongly agree | 171 | 17.0 | 113 | 20.8 | 58 | 12.6 | **<0.001** | 77 | 15.9 | 93 | 18.1 | 0.793 | 142 | 16.6 | 28 | 19.7 | 0.666 | 62 | 14.8 | 104 | 18.9 | **0.007** |
| | Agree | 438 | 43.7 | 242 | 44.6 | 196 | 42.5 | | 220 | 45.5 | 216 | 41.9 | | 376 | 44.0 | 59 | 41.5 | | 174 | 41.4 | 251 | 45.7 | |
| | Neither agree nor disagree | 179 | 17.8 | 77 | 14.2 | 102 | 22.1 | | 93 | 19.3 | 85 | 16.5 | | 153 | 17.9 | 24 | 16.9 | | 83 | 19.8 | 85 | 15.5 | |
| | Disagree | 154 | 15.4 | 78 | 14.4 | 76 | 16.5 | | 62 | 12.8 | 91 | 17.7 | | 132 | 15.4 | 22 | 15.5 | | 71 | 16.9 | 80 | 14.6 | |
| | Strongly disagree | 61 | 6.1 | 32 | 5.9 | 29 | 6.3 | | 31 | 6.4 | 30 | 5.8 | | 52 | 6.1 | 9 | 6.3 | | 30 | 7.1 | 29 | 5.3 | |
| **My cultural or religious traditions make it difficult to follow the guidelines** | Strongly agree | 21 | 2.1 | 15 | 2.8 | 6 | 1.3 | **<0.001** | 12 | 2.5 | 7 | 1.4 | **0.052** | 10 | 1.2 | 11 | 7.7 | **<0.001** | 7 | 1.7 | 13 | 2.4 | 0.245 |
| | Agree | 77 | 7.7 | 61 | 11.3 | 17 | 3.7 | | 42 | 8.7 | 35 | 6.8 | | 42 | 4.9 | 35 | 24.6 | | 30 | 7.1 | 45 | 8.2 | |
| | Neither agree nor disagree | 119 | 11.9 | 71 | 13.1 | 48 | 10.4 | | 65 | 13.5 | 54 | 10.5 | | 97 | 11.3 | 20 | 14.1 | | 46 | 11.0 | 69 | 12.6 | |
| | Disagree | 347 | 34.6 | 186 | 34.3 | 161 | 34.9 | | 162 | 33.5 | 183 | 35.5 | | 294 | 34.4 | 50 | 35.2 | | 146 | 34.8 | 187 | 34.1 | |
| | Strongly disagree | 439 | 43.8 | 209 | 38.6 | 229 | 49.7 | | 202 | 41.8 | 236 | 45.8 | | 412 | 48.2 | 26 | 18.3 | | 191 | 45.5 | 235 | 42.8 | |
| **Cultural food recommendations from healthcare professionals make it difficult to follow the guidelines** | Strongly agree | 13 | 1.3 | 9 | 1.7 | 4 | 0.9 | **<0.001** | 7 | 1.4 | 5 | 1.0 | 0.084 | 5 | 0.6 | 8 | 5.6 | **<0.001** | 4 | 1.0 | 8 | 1.5 | 0.692 |
| | Agree | 76 | 7.6 | 57 | 10.5 | 19 | 4.1 | | 44 | 9.1 | 30 | 5.8 | | 44 | 5.1 | 32 | 22.5 | | 33 | 7.9 | 41 | 7.5 | |
| | Neither agree nor disagree | 180 | 17.9 | 109 | 20.1 | 71 | 15.4 | | 84 | 17.4 | 96 | 18.6 | | 143 | 16.7 | 33 | 23.2 | | 76 | 18.1 | 97 | 17.7 | |
| | Disagree | 332 | 33.1 | 170 | 31.4 | 162 | 35.1 | | 166 | 34.4 | 164 | 31.8 | | 284 | 33.2 | 46 | 32.4 | | 134 | 31.9 | 188 | 34.2 | |
| | Strongly disagree | 402 | 40.1 | 197 | 36.3 | 205 | 44.5 | | 182 | 37.7 | 220 | 42.7 | | 379 | 44.3 | 23 | 16.2 | | 173 | 41.2 | 215 | 39.2 | |
| **My dietary preferences make it difficult to follow the guidelines** | Strongly agree | 26 | 2.6 | 20 | 3.7 | 6 | 1.3 | 0.227 | 6 | 1.2 | 19 | 3.7 | 0.126 | 22 | 2.6 | 3 | 2.1 | **<0.001** | 14 | 3.3 | 11 | 2.0 | 0.482 |
| | Agree | 95 | 9.5 | 63 | 11.6 | 32 | 6.9 | | 40 | 8.3 | 55 | 10.7 | | 69 | 8.1 | 25 | 17.6 | | 44 | 10.5 | 48 | 8.7 | |
| | Neither agree nor disagree | 140 | 14.0 | 69 | 12.7 | 71 | 15.4 | | 65 | 13.5 | 75 | 14.6 | | 108 | 12.6 | 29 | 20.4 | | 66 | 15.7 | 72 | 13.1 | |
| | Disagree | 332 | 33.1 | 169 | 31.2 | 163 | 35.4 | | 171 | 35.4 | 159 | 30.9 | | 291 | 34.0 | 40 | 28.2 | | 123 | 29.3 | 200 | 36.4 | |
| | Strongly disagree | 410 | 40.9 | 221 | 40.8 | 189 | 41.0 | | 201 | 41.6 | 207 | 40.2 | | 365 | 42.7 | 45 | 31.7 | | 173 | 41.2 | 218 | 39.7 | |
| **I struggle to control what I eat** | Strongly agree | 72 | 7.2 | 47 | 8.7 | 25 | 5.4 | **<0.001** | 20 | 4.1 | 52 | 10.1 | **0.005** | 58 | 6.8 | 13 | 9.2 | 0.638 | 7 | 1.7 | 64 | 11.7 | **<0.001** |
| | Agree | 301 | 30.0 | 181 | 33.4 | 120 | 26.0 | | 135 | 28.0 | 161 | 31.3 | | 263 | 30.8 | 36 | 25.4 | | 90 | 21.4 | 203 | 37.0 | |
| | Neither agree nor disagree | 145 | 14.5 | 74 | 13.7 | 71 | 15.4 | | 78 | 16.1 | 67 | 13.0 | | 122 | 14.3 | 22 | 15.5 | | 54 | 12.9 | 89 | 16.2 | |
| | Disagree | 301 | 30.0 | 155 | 28.6 | 146 | 31.7 | | 156 | 32.3 | 145 | 28.2 | | 257 | 30.1 | 42 | 29.6 | | 157 | 37.4 | 134 | 24.4 | |
| | Strongly disagree | 184 | 18.3 | 85 | 15.7 | 99 | 21.5 | | 94 | 19.5 | 90 | 17.5 | | 155 | 18.1 | 29 | 20.4 | | 112 | 26.7 | 59 | 10.7 | |
| **My mood affects what I eat** | Strongly agree | 144 | 14.4 | 98 | 18.1 | 46 | 10.0 | **<0.001** | 50 | 10.4 | 91 | 17.7 | **<0.001** | 115 | 13.5 | 26 | 18.3 | 0.452 | 36 | 8.6 | 106 | 19.3 | **<0.001** |
| | Agree | 389 | 38.8 | 232 | 42.8 | 157 | 34.1 | | 166 | 34.4 | 222 | 43.1 | | 337 | 39.4 | 51 | 35.9 | | 162 | 38.6 | 214 | 39.0 | |

| | | | | | | | | | | | | | | | | | | | | | | | |
|---|---|---|---|---|---|---|---|---|---|---|---|---|---|---|---|---|---|---|---|---|---|---|---|
| | Neither agree nor disagree | 155 | 15.5 | 71 | 13.1 | 84 | 18.2 | | 93 | 19.3 | 62 | 12.0 | | 133 | 15.6 | 20 | 14.1 | | 66 | 15.7 | 83 | 15.1 | |
| | Disagree | 210 | 20.9 | 106 | 19.6 | 104 | 22.6 | | 114 | 23.6 | 95 | 18.4 | | 178 | 20.8 | 32 | 22.5 | | 100 | 23.8 | 105 | 19.1 | |
| | Strongly disagree | 105 | 10.5 | 35 | 6.5 | 70 | 15.2 | | 60 | 12.4 | 45 | 8.7 | | 92 | 10.8 | 13 | 9.2 | | 56 | 13.3 | 41 | 7.5 | |
| **I eat for comfort rather than nutrition** | Strongly agree | 106 | 10.6 | 68 | 12.5 | 38 | 8.2 | **<0.001** | 36 | 7.5 | 68 | 13.2 | **<0.001** | 90 | 10.5 | 15 | 10.6 | 0.809 | 20 | 4.8 | 85 | 15.5 | **<0.001** |
| | Agree | 296 | 29.5 | 180 | 33.2 | 116 | 25.2 | | 129 | 26.7 | 164 | 31.8 | | 249 | 29.1 | 43 | 30.3 | | 81 | 19.3 | 206 | 37.5 | |
| | Neither agree nor disagree | 214 | 21.3 | 105 | 19.4 | 109 | 23.6 | | 111 | 23.0 | 103 | 20.0 | | 181 | 21.2 | 32 | 22.5 | | 104 | 24.8 | 103 | 18.8 | |
| | Disagree | 245 | 24.4 | 124 | 22.9 | 121 | 26.2 | | 127 | 26.3 | 118 | 22.9 | | 215 | 25.1 | 30 | 21.1 | | 128 | 30.5 | 109 | 19.9 | |
| | Strongly disagree | 142 | 14.2 | 65 | 12.0 | 77 | 16.7 | | 80 | 16.6 | 62 | 12.0 | | 120 | 14.0 | 22 | 15.5 | | 87 | 20.7 | 46 | 8.4 | |
| **Eating healthily is too difficult to maintain as part of daily life** | Strongly agree | 43 | 4.3 | 29 | 5.4 | 14 | 3.0 | **0.007** | 15 | 3.1 | 25 | 4.9 | 0.308 | 34 | 4.0 | 9 | 6.3 | 0.102 | 13 | 3.1 | 29 | 5.3 | **<0.001** |
| | Agree | 238 | 23.7 | 155 | 28.6 | 83 | 18.0 | | 104 | 21.5 | 132 | 25.6 | | 195 | 22.8 | 41 | 28.9 | | 77 | 18.3 | 151 | 27.5 | |
| | Neither agree nor disagree | 279 | 27.8 | 124 | 22.9 | 155 | 33.6 | | 148 | 30.6 | 131 | 25.4 | | 239 | 28.0 | 37 | 26.1 | | 100 | 23.8 | 174 | 31.7 | |
| | Disagree | 330 | 32.9 | 178 | 32.8 | 152 | 33.0 | | 163 | 33.7 | 167 | 32.4 | | 292 | 34.2 | 37 | 26.1 | | 165 | 39.3 | 151 | 27.5 | |
| | Strongly disagree | 113 | 11.3 | 56 | 10.3 | 57 | 12.4 | | 53 | 11.0 | 60 | 11.7 | | 95 | 11.1 | 18 | 12.7 | | 65 | 15.5 | 44 | 8.0 | |
| **I don't need to follow the guidelines because I take vitamin supplements** | Strongly agree | 11 | 1.1 | 10 | 1.8 | 1 | 0.2 | **0.009** | 8 | 1.7 | 3 | 0.6 | 0.200 | 7 | 0.8 | 4 | 2.8 | **0.031** | 6 | 1.4 | 5 | 0.9 | 0.847 |
| | Agree | 39 | 3.9 | 27 | 5.0 | 12 | 2.6 | | 21 | 4.3 | 18 | 3.5 | | 28 | 3.3 | 11 | 7.7 | | 18 | 4.3 | 18 | 3.3 | |
| | Neither agree nor disagree | 76 | 7.6 | 48 | 8.9 | 28 | 6.1 | | 43 | 8.9 | 33 | 6.4 | | 61 | 7.1 | 13 | 9.2 | | 29 | 6.9 | 42 | 7.7 | |
| | Disagree | 455 | 45.4 | 242 | 44.6 | 213 | 46.2 | | 212 | 43.9 | 239 | 46.4 | | 391 | 45.7 | 61 | 43.0 | | 189 | 45.0 | 251 | 45.7 | |
| | Strongly disagree | 422 | 42.1 | 215 | 39.7 | 207 | 44.9 | | 199 | 41.2 | 222 | 43.1 | | 368 | 43.0 | 53 | 37.3 | | 178 | 42.4 | 233 | 42.4 | |

Note. n = 1003. Age: Under 50 (n=542) vs 50 and over (n=461). Sex: Male (n=483) vs Female (n=515). Ethnicity: White (n=855) vs Non-white (n=142). BMI: Healthy weight (n=420) vs Overweight/obesity (n=549). Bold p values indicate statistical significance ($p<0.05$). p values from Mann-Whitney U test.

**Supplementary Table 7. Response distributions and group comparisons for perceived facilitators to following UK Government healthy eating guidelines. p values from Mann-Whitney U test.**

| Question | Response | Overall (n=1003) | | Age | | | | | Sex | | | | | Ethnicity | | | | | BMI | | | | |
|---|---|---|---|---|---|---|---|---|---|---|---|---|---|---|---|---|---|---|---|---|---|---|---|
| | | n | % | Under 50 | | 50 and over | | p | Male | | Female | | p | White | | Non-white | | p | Healthy weight | | Overweight/ obesity | | p |
| | | | | n | % | n | % | | n | % | n | % | | n | % | n | % | | n | % | n | % | |
| **Having more opportunities to learn about the guidelines would help me follow them** | Strongly agree | 85 | 8.5 | 52 | 9.6 | 33 | 7.2 | **0.025** | 36 | 7.5 | 49 | 9.5 | 0.924 | 62 | 7.3 | 22 | 15.5 | **<0.001** | 31 | 7.4 | 51 | 9.3 | 0.932 |
| | Agree | 413 | 41.2 | 240 | 44.3 | 173 | 37.5 | | 211 | 43.7 | 200 | 38.8 | | 339 | 39.6 | 72 | 50.7 | | 183 | 43.6 | 220 | 40.1 | |
| | Neither agree nor disagree | 282 | 28.1 | 130 | 24.0 | 152 | 33.0 | | 126 | 26.1 | 156 | 30.3 | | 252 | 29.5 | 28 | 19.7 | | 115 | 27.4 | 157 | 28.6 | |
| | Disagree | 165 | 16.5 | 90 | 16.6 | 75 | 16.3 | | 83 | 17.2 | 80 | 15.5 | | 148 | 17.3 | 16 | 11.3 | | 67 | 16.0 | 90 | 16.4 | |
| | Strongly disagree | 58 | 5.8 | 30 | 5.5 | 28 | 6.1 | | 27 | 5.6 | 30 | 5.8 | | 54 | 6.3 | 4 | 2.8 | | 24 | 5.7 | 31 | 5.6 | |
| **Greater public health promotion of the guidelines would help me follow them** | Strongly agree | 122 | 12.2 | 82 | 15.1 | 40 | 8.7 | **0.003** | 55 | 11.4 | 67 | 13.0 | 0.794 | 95 | 11.1 | 26 | 18.3 | **<0.001** | 53 | 12.6 | 68 | 12.4 | 0.718 |
| | Agree | 413 | 41.2 | 225 | 41.5 | 190 | 41.2 | | 212 | 43.9 | 201 | 39.0 | | 347 | 40.6 | 67 | 47.2 | | 181 | 43.1 | 224 | 40.8 | |
| | Neither agree nor disagree | 215 | 21.4 | 111 | 20.5 | 104 | 22.6 | | 94 | 19.5 | 120 | 23.3 | | 183 | 21.4 | 28 | 19.7 | | 79 | 18.8 | 126 | 23.0 | |
| | Disagree | 175 | 17.4 | 93 | 17.2 | 84 | 18.2 | | 86 | 17.8 | 89 | 17.3 | | 161 | 18.8 | 16 | 11.3 | | 79 | 18.8 | 90 | 16.4 | |
| | Strongly disagree | 78 | 7.8 | 31 | 5.7 | 43 | 9.3 | | 36 | 7.5 | 38 | 7.4 | | 69 | 8.1 | 5 | 3.5 | | 28 | 6.7 | 41 | 7.5 | |
| **Being able to find the Eatwell Guide easily would help me follow it** | Strongly agree | 134 | 13.4 | 79 | 14.6 | 55 | 11.9 | 0.151 | 61 | 12.6 | 73 | 14.2 | 0.692 | 108 | 12.6 | 25 | 17.6 | **<0.001** | 56 | 13.3 | 75 | 13.7 | 0.612 |
| | Agree | 362 | 36.1 | 201 | 37.1 | 161 | 34.9 | | 185 | 38.3 | 175 | 34.0 | | 291 | 34.0 | 69 | 48.6 | | 159 | 37.9 | 192 | 35.0 | |
| | Neither agree nor disagree | 225 | 22.4 | 114 | 21.0 | 111 | 24.1 | | 106 | 21.9 | 119 | 23.1 | | 201 | 23.5 | 23 | 16.2 | | 92 | 21.9 | 127 | 23.1 | |
| | Disagree | 199 | 19.8 | 105 | 19.4 | 94 | 20.4 | | 90 | 18.6 | 107 | 20.8 | | 180 | 21.1 | 18 | 12.7 | | 79 | 18.8 | 112 | 20.4 | |
| | Strongly disagree | 83 | 8.3 | 43 | 7.9 | 40 | 8.7 | | 41 | 8.5 | 41 | 8.0 | | 75 | 8.8 | 7 | 4.9 | | 34 | 8.1 | 43 | 7.8 | |
| **Making the Eatwell Guide easier to understand would help me follow it** | Strongly agree | 72 | 7.2 | 52 | 9.6 | 20 | 4.3 | **<0.001** | 29 | 6.0 | 43 | 8.3 | 0.940 | 49 | 5.7 | 23 | 16.2 | **<0.001** | 34 | 8.1 | 36 | 6.6 | 0.510 |
| | Agree | 224 | 22.3 | 145 | 26.8 | 81 | 17.6 | | 112 | 23.2 | 112 | 21.7 | | 171 | 20.0 | 54 | 38.0 | | 94 | 22.4 | 126 | 23.0 | |
| | Neither agree nor disagree | 363 | 36.2 | 180 | 33.2 | 183 | 39.7 | | 180 | 37.3 | 181 | 35.1 | | 324 | 37.9 | 35 | 24.6 | | 158 | 37.6 | 193 | 35.2 | |
| | Disagree | 252 | 25.1 | 122 | 22.5 | 130 | 28.2 | | 120 | 24.8 | 131 | 25.4 | | 229 | 26.8 | 22 | 15.5 | | 91 | 21.7 | 150 | 27.3 | |
| | Strongly disagree | 92 | 9.2 | 43 | 7.9 | 47 | 10.2 | | 42 | 8.7 | 48 | 9.3 | | 82 | 9.6 | 8 | 5.6 | | 43 | 10.2 | 44 | 8.0 | |
| **Having more budget-friendly healthy meal ideas would help me follow the guidelines** | Strongly agree | 277 | 27.6 | 193 | 35.6 | 84 | 18.2 | **<0.001** | 117 | 24.2 | 160 | 31.1 | **0.034** | 216 | 25.3 | 59 | 41.5 | **0.001** | 121 | 28.8 | 151 | 27.5 | 0.916 |
| | Agree | 446 | 44.5 | 231 | 42.6 | 215 | 46.6 | | 224 | 46.4 | 218 | 42.3 | | 393 | 46.0 | 52 | 36.6 | | 179 | 42.6 | 250 | 45.5 | |
| | Neither agree nor disagree | 170 | 16.9 | 64 | 11.8 | 106 | 23.0 | | 84 | 17.4 | 86 | 16.7 | | 151 | 17.7 | 18 | 12.7 | | 74 | 17.6 | 93 | 16.9 | |
| | Disagree | 83 | 8.3 | 43 | 7.9 | 40 | 8.7 | | 42 | 8.7 | 40 | 7.8 | | 68 | 8.0 | 13 | 9.2 | | 34 | 8.1 | 44 | 8.0 | |
| | Strongly disagree | 27 | 2.7 | 11 | 2.0 | 16 | 3.5 | | 16 | 3.3 | 11 | 2.1 | | 27 | 3.2 | 0 | 0.0 | | 12 | 2.9 | 11 | 2.0 | |
| **Having guidelines** | Strongly agree | 179 | 17.8 | 118 | 21.8 | 62 | 13.4 | **<0.001** | 73 | 15.1 | 106 | 20.6 | **0.005** | 145 | 17.0 | 34 | 23.9 | **0.011** | 81 | 19.3 | 94 | 17.1 | 0.430 |

| | | | | | | | | | | | | | | | | | | | | | | | |
|---|---|---|---|---|---|---|---|---|---|---|---|---|---|---|---|---|---|---|---|---|---|---|---|
| **tailored to my specific health needs or requirements would help** | Agree | 366 | 36.5 | 210 | 38.7 | 156 | 33.8 | | 167 | 34.6 | 197 | 38.3 | | 307 | 35.9 | 55 | 38.7 | | 152 | 36.2 | 204 | 37.2 | |
| | Neither agree nor disagree | 277 | 27.6 | 128 | 23.6 | 150 | 32.5 | | 152 | 31.5 | 125 | 24.3 | | 243 | 28.4 | 34 | 23.9 | | 118 | 28.1 | 152 | 27.7 | |
| | Disagree | 130 | 13.0 | 63 | 11.6 | 67 | 14.5 | | 65 | 13.5 | 65 | 12.6 | | 114 | 13.3 | 16 | 11.3 | | 53 | 12.6 | 71 | 12.9 | |
| | Strongly disagree | 51 | 5.1 | 23 | 4.2 | 26 | 5.6 | | 26 | 5.4 | 22 | 4.3 | | 46 | 5.4 | 3 | 2.1 | | 16 | 3.8 | 28 | 5.1 | |
| **Learning how to cook affordable healthy meals would help me follow the guidelines** | Strongly agree | 172 | 17.1 | 123 | 22.7 | 49 | 10.6 | **<0.001** | 77 | 15.9 | 95 | 18.4 | 0.612 | 137 | 16.0 | 34 | 23.9 | **0.001** | 71 | 16.9 | 95 | 17.3 | 0.872 |
| | Agree | 351 | 35.0 | 202 | 37.3 | 151 | 32.8 | | 175 | 36.2 | 176 | 34.2 | | 293 | 34.3 | 57 | 40.1 | | 147 | 35.0 | 194 | 35.3 | |
| | Neither agree nor disagree | 259 | 25.8 | 117 | 21.6 | 143 | 31.0 | | 125 | 25.9 | 134 | 26.0 | | 228 | 26.7 | 31 | 21.8 | | 112 | 26.7 | 141 | 25.7 | |
| | Disagree | 150 | 15.0 | 70 | 12.9 | 81 | 17.6 | | 74 | 15.3 | 75 | 14.6 | | 137 | 16.0 | 13 | 9.2 | | 62 | 14.8 | 83 | 15.1 | |
| | Strongly disagree | 71 | 7.1 | 30 | 5.5 | 37 | 8.0 | | 32 | 6.6 | 35 | 6.8 | | 60 | 7.0 | 7 | 4.9 | | 28 | 6.7 | 36 | 6.6 | |
| **Having more examples of what healthy portion sizes look like would help me** | Strongly agree | 163 | 16.3 | 117 | 21.6 | 47 | 10.2 | **<0.001** | 64 | 13.3 | 99 | 19.2 | **0.039** | 128 | 15.0 | 35 | 24.6 | **<0.001** | 64 | 15.2 | 94 | 17.1 | 0.272 |
| | Agree | 453 | 45.2 | 248 | 45.8 | 205 | 44.5 | | 221 | 45.8 | 230 | 44.7 | | 379 | 44.3 | 71 | 50.0 | | 189 | 45.0 | 248 | 45.2 | |
| | Neither agree nor disagree | 199 | 19.8 | 89 | 16.4 | 110 | 23.9 | | 105 | 21.7 | 92 | 17.9 | | 174 | 20.4 | 24 | 16.9 | | 83 | 19.8 | 109 | 19.9 | |
| | Disagree | 126 | 12.6 | 58 | 10.7 | 68 | 14.8 | | 64 | 13.3 | 62 | 12.0 | | 117 | 13.7 | 8 | 5.6 | | 50 | 11.9 | 74 | 13.5 | |
| | Strongly disagree | 62 | 6.2 | 30 | 5.5 | 31 | 6.7 | | 29 | 6.0 | 32 | 6.2 | | 57 | 6.7 | 4 | 2.8 | | 34 | 8.1 | 24 | 4.4 | |
| **Having more culturally appropriate food options in the guidelines would help me** | Strongly agree | 88 | 8.8 | 70 | 12.9 | 18 | 3.9 | **<0.001** | 35 | 7.2 | 53 | 10.3 | 0.931 | 57 | 6.7 | 31 | 21.8 | **<0.001** | 30 | 7.1 | 56 | 10.2 | 0.734 |
| | Agree | 242 | 24.1 | 147 | 27.1 | 95 | 20.6 | | 130 | 26.9 | 110 | 21.4 | | 185 | 21.6 | 53 | 37.3 | | 109 | 26.0 | 126 | 23.0 | |
| | Neither agree nor disagree | 288 | 28.7 | 141 | 26.0 | 147 | 31.9 | | 130 | 26.9 | 157 | 30.5 | | 253 | 29.6 | 34 | 23.9 | | 120 | 28.6 | 157 | 28.6 | |
| | Disagree | 222 | 22.1 | 99 | 18.3 | 123 | 26.7 | | 113 | 23.4 | 108 | 21.0 | | 206 | 24.1 | 16 | 11.3 | | 92 | 21.9 | 122 | 22.2 | |
| | Strongly disagree | 163 | 16.3 | 85 | 15.7 | 78 | 16.9 | | 75 | 15.5 | 87 | 16.9 | | 154 | 18.0 | 8 | 5.6 | | 69 | 16.4 | 88 | 16.0 | |
| **Receiving personalised dietary advice from a healthcare professional would help me** | Strongly agree | 168 | 16.8 | 133 | 24.5 | 37 | 8.0 | **<0.001** | 65 | 13.5 | 104 | 20.2 | **0.052** | 125 | 14.6 | 43 | 30.3 | **<0.001** | 68 | 16.2 | 97 | 17.7 | 0.330 |
| | Agree | 336 | 33.5 | 212 | 39.1 | 127 | 27.5 | | 172 | 35.6 | 163 | 31.7 | | 282 | 33.0 | 54 | 38.0 | | 139 | 33.1 | 187 | 34.1 | |
| | Neither agree nor disagree | 245 | 24.4 | 98 | 18.1 | 147 | 31.9 | | 115 | 23.8 | 130 | 25.2 | | 218 | 25.5 | 27 | 19.0 | | 103 | 24.5 | 135 | 24.6 | |
| | Disagree | 169 | 16.9 | 67 | 12.4 | 102 | 22.1 | | 91 | 18.8 | 78 | 15.1 | | 155 | 18.1 | 13 | 9.2 | | 74 | 17.6 | 90 | 16.4 | |
| | Strongly disagree | 85 | 8.5 | 32 | 5.9 | 48 | 10.4 | | 40 | 8.3 | 40 | 7.8 | | 75 | 8.8 | 5 | 3.5 | | 36 | 8.6 | 40 | 7.3 | |
| **Having healthier food options available at work would help me follow the guidelines** | Strongly agree | 125 | 12.5 | 96 | 17.7 | 29 | 6.3 | **<0.001** | 41 | 8.5 | 84 | 16.3 | 0.455 | 91 | 10.6 | 33 | 23.2 | **<0.001** | 57 | 13.6 | 63 | 11.5 | 0.125 |
| | Agree | 289 | 28.8 | 196 | 36.2 | 93 | 20.2 | | 160 | 33.1 | 127 | 24.7 | | 235 | 27.5 | 53 | 37.3 | | 127 | 30.2 | 153 | 27.9 | |
| | Neither agree nor disagree | 312 | 31.1 | 134 | 24.7 | 178 | 38.6 | | 151 | 31.3 | 160 | 31.1 | | 282 | 33.0 | 28 | 19.7 | | 130 | 31.0 | 172 | 31.3 | |
| | Disagree | 161 | 16.1 | 71 | 13.1 | 90 | 19.5 | | 74 | 15.3 | 87 | 16.9 | | 142 | 16.6 | 18 | 12.7 | | 59 | 14.0 | 98 | 17.9 | |
| | Strongly disagree | 116 | 11.6 | 45 | 8.3 | 71 | 15.4 | | 57 | 11.8 | 57 | 11.1 | | 105 | 12.3 | 10 | 7.0 | | 47 | 11.2 | 63 | 11.5 | |
| **Having face-to-face support from** | Strongly agree | 107 | 10.7 | 66 | 12.2 | 41 | 8.9 | **0.017** | 44 | 9.1 | 63 | 12.2 | 0.474 | 90 | 10.5 | 15 | 10.6 | **0.015** | 39 | 9.3 | 67 | 12.2 | 0.177 |
| | Agree | 300 | 29.9 | 171 | 31.5 | 129 | 28.0 | | 150 | 31.1 | 149 | 28.9 | | 247 | 28.9 | 53 | 37.3 | | 116 | 27.6 | 171 | 31.1 | |

| | | | | | | | | | | | | | | | | | | | | | |
|---|---|---|---|---|---|---|---|---|---|---|---|---|---|---|---|---|---|---|---|---|---|
| **a professional would help me follow the guidelines** | Neither agree nor disagree | 252 | 25.1 | 136 | 25.1 | 116 | 25.2 | | 124 | 25.7 | 127 | 24.7 | | 209 | 24.4 | 42 | 29.6 | | 121 | 28.8 | 123 | 22.4 | |
| | Disagree | 228 | 22.7 | 108 | 19.9 | 121 | 26.2 | | 105 | 21.7 | 123 | 23.9 | | 207 | 24.2 | 20 | 14.1 | | 96 | 22.9 | 128 | 23.3 | |
| | Strongly disagree | 116 | 11.6 | 61 | 11.3 | 54 | 11.7 | | 60 | 12.4 | 53 | 10.3 | | 102 | 11.9 | 12 | 8.5 | | 48 | 11.4 | 60 | 10.9 | |
| **Having access to an app or website to help me follow the guidelines would help** | Strongly agree | 124 | 12.4 | 78 | 14.4 | 46 | 10.0 | **0.017** | 42 | 8.7 | 82 | 15.9 | 0.104 | 100 | 11.7 | 24 | 16.9 | **0.018** | 46 | 11.0 | 77 | 14.0 | 0.276 |
| | Agree | 415 | 41.4 | 227 | 41.9 | 188 | 40.8 | | 211 | 43.7 | 204 | 39.6 | | 347 | 40.6 | 64 | 45.1 | | 174 | 41.4 | 226 | 41.2 | |
| | Neither agree nor disagree | 241 | 24.0 | 128 | 23.6 | 113 | 24.5 | | 123 | 25.5 | 115 | 22.3 | | 209 | 24.4 | 31 | 21.8 | | 108 | 25.7 | 124 | 22.6 | |
| | Disagree | 148 | 14.8 | 75 | 13.8 | 74 | 16.1 | | 75 | 15.5 | 73 | 14.2 | | 136 | 15.9 | 12 | 8.5 | | 56 | 13.3 | 88 | 16.0 | |
| | Strongly disagree | 75 | 7.5 | 34 | 6.3 | 40 | 8.7 | | 32 | 6.6 | 41 | 8.0 | | 63 | 7.4 | 11 | 7.7 | | 36 | 8.6 | 34 | 6.2 | |
| **If healthy foods were cheaper I would be more likely to follow the guidelines** | Strongly agree | 369 | 36.8 | 234 | 43.2 | 135 | 29.3 | **<0.001** | 161 | 33.3 | 206 | 40.0 | **0.098** | 310 | 36.3 | 57 | 40.1 | 0.086 | 145 | 34.5 | 213 | 38.8 | 0.332 |
| | Agree | 373 | 37.2 | 201 | 37.1 | 172 | 37.3 | | 193 | 40.0 | 178 | 34.6 | | 313 | 36.6 | 58 | 40.8 | | 163 | 38.8 | 197 | 35.9 | |
| | Neither agree nor disagree | 147 | 14.7 | 60 | 11.1 | 87 | 18.9 | | 74 | 15.3 | 72 | 14.0 | | 128 | 15.0 | 17 | 12.0 | | 70 | 16.7 | 73 | 13.3 | |
| | Disagree | 80 | 8.0 | 34 | 6.3 | 46 | 10.0 | | 35 | 7.2 | 45 | 8.7 | | 72 | 8.4 | 8 | 5.6 | | 28 | 6.7 | 49 | 8.9 | |
| | Strongly disagree | 34 | 3.4 | 13 | 2.4 | 21 | 4.6 | | 20 | 4.1 | 14 | 2.7 | | 32 | 3.7 | 2 | 1.4 | | 14 | 3.3 | 17 | 3.1 | |
| **Support from family or friends would help me follow the guidelines** | Strongly agree | 120 | 12.0 | 87 | 16.1 | 33 | 7.2 | **<0.001** | 53 | 11.0 | 67 | 13.0 | 0.356 | 96 | 11.2 | 23 | 16.2 | **<0.001** | 44 | 10.5 | 72 | 13.1 | 0.303 |
| | Agree | 329 | 32.8 | 205 | 37.8 | 124 | 26.9 | | 158 | 32.7 | 170 | 33.0 | | 261 | 30.5 | 66 | 46.5 | | 135 | 32.1 | 186 | 33.9 | |
| | Neither agree nor disagree | 328 | 32.7 | 159 | 29.3 | 169 | 36.7 | | 160 | 33.1 | 167 | 32.4 | | 288 | 33.7 | 38 | 26.8 | | 150 | 35.7 | 165 | 30.1 | |
| | Disagree | 159 | 15.9 | 63 | 11.6 | 98 | 21.3 | | 81 | 16.8 | 78 | 15.1 | | 150 | 17.5 | 11 | 7.7 | | 63 | 15.0 | 96 | 17.5 | |
| | Strongly disagree | 67 | 6.7 | 28 | 5.2 | 37 | 8.0 | | 31 | 6.4 | 33 | 6.4 | | 60 | 7.0 | 4 | 2.8 | | 28 | 6.7 | 30 | 5.5 | |
| **If restaurants and takeaways offered healthier options this would help me** | Strongly agree | 211 | 21.0 | 132 | 24.4 | 79 | 17.1 | **0.002** | 85 | 17.6 | 126 | 24.5 | **0.044** | 177 | 20.7 | 33 | 23.2 | **0.039** | 90 | 21.4 | 116 | 21.1 | 0.528 |
| | Agree | 394 | 39.3 | 214 | 39.5 | 180 | 39.0 | | 201 | 41.6 | 191 | 37.1 | | 327 | 38.2 | 65 | 45.8 | | 153 | 36.4 | 225 | 41.0 | |
| | Neither agree nor disagree | 209 | 20.8 | 103 | 19.0 | 106 | 23.0 | | 98 | 20.3 | 111 | 21.6 | | 180 | 21.1 | 27 | 19.0 | | 99 | 23.6 | 106 | 19.3 | |
| | Disagree | 130 | 13.0 | 67 | 12.4 | 63 | 13.7 | | 68 | 14.1 | 62 | 12.0 | | 121 | 14.2 | 9 | 6.3 | | 56 | 13.3 | 70 | 12.8 | |
| | Strongly disagree | 59 | 5.9 | 26 | 4.8 | 33 | 7.2 | | 31 | 6.4 | 25 | 4.9 | | 50 | 5.8 | 8 | 5.6 | | 22 | 5.2 | 32 | 5.8 | |
| **Having calorie information on menus would help me follow the guidelines** | Strongly agree | 173 | 17.2 | 114 | 21.0 | 59 | 12.8 | **0.014** | 77 | 15.9 | 96 | 18.6 | 0.781 | 150 | 17.5 | 22 | 15.5 | 0.104 | 66 | 15.7 | 103 | 18.8 | **0.047** |
| | Agree | 366 | 36.5 | 193 | 35.6 | 173 | 37.5 | | 189 | 39.1 | 176 | 34.2 | | 296 | 34.6 | 67 | 47.2 | | 147 | 35.0 | 210 | 38.3 | |
| | Neither agree nor disagree | 204 | 20.3 | 99 | 18.3 | 105 | 22.8 | | 90 | 18.6 | 114 | 22.1 | | 178 | 20.8 | 25 | 17.6 | | 91 | 21.7 | 106 | 19.3 | |
| | Disagree | 176 | 17.5 | 91 | 16.8 | 86 | 18.7 | | 85 | 17.6 | 91 | 17.7 | | 158 | 18.5 | 19 | 13.4 | | 79 | 18.8 | 91 | 16.6 | |
| | Strongly disagree | 84 | 8.4 | 45 | 8.3 | 38 | 8.2 | | 42 | 8.7 | 38 | 7.4 | | 73 | 8.5 | 9 | 6.3 | | 37 | 8.8 | 39 | 7.1 | |
| **Having healthy meal box delivery** | Strongly agree | 117 | 11.7 | 80 | 14.8 | 37 | 8.0 | **<0.001** | 49 | 10.1 | 68 | 13.2 | 0.283 | 103 | 12.0 | 13 | 9.2 | 0.233 | 49 | 11.7 | 66 | 12.0 | 0.909 |
| | Agree | 324 | 32.3 | 196 | 36.2 | 129 | 28.0 | | 156 | 32.3 | 168 | 32.6 | | 265 | 31.0 | 58 | 40.8 | | 141 | 33.6 | 173 | 31.5 | |

| | | | | | | | | | | | | | | | | | | | | | |
|---|---|---|---|---|---|---|---|---|---|---|---|---|---|---|---|---|---|---|---|---|---|
| **options would help me follow the guidelines** | Neither agree nor disagree | 283 | 28.2 | 135 | 24.9 | 148 | 32.1 | | 140 | 29.0 | 143 | 27.8 | | 242 | 28.3 | 40 | 28.2 | | 111 | 26.4 | 163 | 29.7 | |
| | Disagree | 162 | 16.2 | 78 | 14.4 | 85 | 18.4 | | 89 | 18.4 | 73 | 14.2 | | 147 | 17.2 | 15 | 10.6 | | 67 | 16.0 | 91 | 16.6 | |
| | Strongly disagree | 117 | 11.7 | 53 | 9.8 | 62 | 13.4 | | 49 | 10.1 | 63 | 12.2 | | 98 | 11.5 | 16 | 11.3 | | 52 | 12.4 | 56 | 10.2 | |
| **If following the guidelines would improve my health, I would be more likely to follow them** | Strongly agree | 195 | 19.4 | 128 | 23.6 | 67 | 14.5 | **0.001** | 90 | 18.6 | 103 | 20.0 | 0.712 | 161 | 18.8 | 32 | 22.5 | **0.012** | 84 | 20.0 | 102 | 18.6 | 0.419 |
| | Agree | 440 | 43.9 | 238 | 43.9 | 202 | 43.8 | | 217 | 44.9 | 221 | 42.9 | | 364 | 42.6 | 74 | 52.1 | | 191 | 45.5 | 238 | 43.4 | |
| | Neither agree nor disagree | 244 | 24.3 | 108 | 19.9 | 136 | 29.5 | | 111 | 23.0 | 133 | 25.8 | | 223 | 26.1 | 21 | 14.8 | | 92 | 21.9 | 145 | 26.4 | |
| | Disagree | 87 | 8.7 | 50 | 9.2 | 37 | 8.0 | | 45 | 9.3 | 42 | 8.2 | | 73 | 8.5 | 13 | 9.2 | | 35 | 8.3 | 48 | 8.7 | |
| | Strongly disagree | 37 | 3.7 | 18 | 3.3 | 19 | 4.1 | | 20 | 4.1 | 16 | 3.1 | | 34 | 4.0 | 2 | 1.4 | | 18 | 4.3 | 16 | 2.9 | |
| **If following the guidelines would improve my mental health, I would be more likely to follow them** | Strongly agree | 183 | 18.2 | 127 | 23.4 | 56 | 12.1 | **<0.001** | 81 | 16.8 | 100 | 19.4 | 0.806 | 153 | 17.9 | 29 | 20.4 | **0.048** | 84 | 20.0 | 95 | 17.3 | **0.024** |
| | Agree | 473 | 47.2 | 260 | 48.0 | 213 | 46.2 | | 245 | 50.7 | 226 | 43.9 | | 395 | 46.2 | 75 | 52.8 | | 209 | 49.8 | 249 | 45.4 | |
| | Neither agree nor disagree | 207 | 20.6 | 94 | 17.3 | 113 | 24.5 | | 88 | 18.2 | 119 | 23.1 | | 182 | 21.3 | 25 | 17.6 | | 79 | 18.8 | 120 | 21.9 | |
| | Disagree | 100 | 10.0 | 48 | 8.9 | 52 | 11.3 | | 49 | 10.1 | 51 | 9.9 | | 90 | 10.5 | 9 | 6.3 | | 34 | 8.1 | 63 | 11.5 | |
| | Strongly disagree | 40 | 4.0 | 13 | 2.4 | 27 | 5.9 | | 20 | 4.1 | 19 | 3.7 | | 35 | 4.1 | 4 | 2.8 | | 14 | 3.3 | 22 | 4.0 | |
| **If following the guidelines would help me manage my weight, I would be more likely to follow them** | Strongly agree | 205 | 20.4 | 137 | 25.3 | 69 | 15.0 | **<0.001** | 80 | 16.6 | 126 | 24.5 | **0.078** | 164 | 19.2 | 41 | 28.9 | **0.009** | 77 | 18.3 | 124 | 22.6 | **0.033** |
| | Agree | 451 | 45.0 | 248 | 45.8 | 206 | 44.7 | | 237 | 49.1 | 213 | 41.4 | | 389 | 45.5 | 62 | 43.7 | | 190 | 45.2 | 252 | 45.9 | |
| | Neither agree nor disagree | 219 | 21.8 | 98 | 18.1 | 121 | 26.2 | | 101 | 20.9 | 118 | 22.9 | | 193 | 22.6 | 26 | 18.3 | | 95 | 22.6 | 115 | 20.9 | |
| | Disagree | 84 | 8.4 | 38 | 7.0 | 47 | 10.2 | | 45 | 9.3 | 40 | 7.8 | | 76 | 8.9 | 8 | 5.6 | | 35 | 8.3 | 47 | 8.6 | |
| | Strongly disagree | 44 | 4.4 | 21 | 3.9 | 18 | 3.9 | | 20 | 4.1 | 18 | 3.5 | | 33 | 3.9 | 5 | 3.5 | | 23 | 5.5 | 11 | 2.0 | |
| **If healthy food was more enjoyable I would be more likely to follow the guidelines** | Strongly agree | 183 | 18.2 | 101 | 18.6 | 82 | 17.8 | 0.712 | 75 | 15.5 | 108 | 21.0 | **0.013** | 148 | 17.3 | 35 | 24.6 | **0.002** | 91 | 21.7 | 87 | 15.8 | **<0.001** |
| | Agree | 435 | 43.4 | 240 | 44.3 | 195 | 42.3 | | 203 | 42.0 | 230 | 44.7 | | 363 | 42.5 | 69 | 48.6 | | 203 | 48.3 | 214 | 39.0 | |
| | Neither agree nor disagree | 216 | 21.5 | 105 | 19.4 | 111 | 24.1 | | 124 | 25.7 | 90 | 17.5 | | 193 | 22.6 | 21 | 14.8 | | 73 | 17.4 | 134 | 24.4 | |
| | Disagree | 129 | 12.9 | 68 | 12.5 | 61 | 13.2 | | 61 | 12.6 | 68 | 13.2 | | 118 | 13.8 | 10 | 7.0 | | 41 | 9.8 | 88 | 16.0 | |
| | Strongly disagree | 40 | 4.0 | 28 | 5.2 | 12 | 2.6 | | 20 | 4.1 | 19 | 3.7 | | 33 | 3.9 | 7 | 4.9 | | 12 | 2.9 | 26 | 4.7 | |
| **I would be more likely to follow the guidelines if I was more motivated to do so** | Strongly agree | 105 | 10.5 | 62 | 11.4 | 43 | 9.3 | 0.758 | 42 | 8.7 | 63 | 12.2 | 0.162 | 88 | 10.3 | 17 | 12.0 | 0.501 | 43 | 10.2 | 58 | 10.6 | **0.014** |
| | Agree | 413 | 41.2 | 221 | 40.8 | 192 | 41.6 | | 197 | 40.8 | 214 | 41.6 | | 352 | 41.2 | 58 | 40.8 | | 196 | 46.7 | 206 | 37.5 | |
| | Neither agree nor disagree | 315 | 31.4 | 163 | 30.1 | 152 | 33.0 | | 164 | 34.0 | 150 | 29.1 | | 266 | 31.1 | 47 | 33.1 | | 123 | 29.3 | 179 | 32.6 | |
| | Disagree | 127 | 12.7 | 70 | 12.9 | 57 | 12.4 | | 58 | 12.0 | 68 | 13.2 | | 112 | 13.1 | 14 | 9.9 | | 41 | 9.8 | 82 | 14.9 | |
| | Strongly disagree | 43 | 4.3 | 26 | 4.8 | 17 | 3.7 | | 22 | 4.6 | 20 | 3.9 | | 37 | 4.3 | 6 | 4.2 | | 17 | 4.0 | 24 | 4.4 | |

Note. n = 1003. Age: Under 50 (n=542) vs 50 and over (n=461). Sex: Male (n=483) vs Female (n=515). Ethnicity: White (n=855) vs Non-white (n=142). BMI: Healthy weight (n=420) vs Overweight/obesity (n=549). Bold p values indicate statistical significance (p<0.05).

**Supplementary table 8. Subgroup comparison of knowledge scores**

| Knowledge score | Age | | | Sex | | | Ethnicity | | | BMI | | |
|---|---|---|---|---|---|---|---|---|---|---|---|---|
| | Under 50 (n=542) | 50 and over (n=461) | p | Male (n=483) | Female (n=515) | p | White (n=855) | Non-white (n=142) | p | Healthy weight (n=420) | Overweight/ obesity (n=549) | p |
| **EWG knowledge (strict), mean % (SD)** | 54.3 (15.3) | 52.1 (15.4) | **0.019** | 51.5 (15.9) | 55.0 (14.8) | **<0.001** | 53.9 (15.1) | 49.6 (17.0) | **0.004** | 53.3 (14.7) | 53.4 (15.5) | 0.931 |
| **EWG knowledge (liberal), mean % (SD)** | 71.8 (13.8) | 73.3 (14.2) | **0.023** | 70.7 (15.0) | 74.1 (12.9) | **<0.001** | 73.3 (13.7) | 67.9 (14.8) | **<0.001** | 71.9 (13.7) | 73.1 (13.8) | 0.148 |
| **Broader knowledge (strict), mean % (SD)** | 20.5 (16.7) | 14.9 (15.5) | **<0.001** | 18.3 (16.8) | 17.5 (16.1) | 0.455 | 17.8 (15.8) | 18.9 (19.8) | 0.892 | 18.2 (15.5) | 17.7 (17.0) | 0.221 |
| **Broader knowledge (liberal), mean % (SD)** | 61.2 (18.6) | 56.2 (19.6) | **<0.001** | 60.1 (19.6) | 57.8 (18.8) | **0.017** | 58.9 (18.9) | 59.2 (20.9) | 0.828 | 60.3 (18.6) | 57.9 (19.4) | 0.081 |
